# Association between Maternal Depression during Pregnancy and Newborn DNA Methylation

**DOI:** 10.1101/2021.06.02.21258194

**Authors:** Emily Drzymalla, Nicole Gladish, Nastassja Koen, Michael P. Epstein, Michael S. Kobor, Heather J. Zar, Dan J. Stein, Anke Huels

## Abstract

Around 15% to 65% of women globally experience depression during pregnancy, prevalence being particularly high in low- and middle-income countries. Prenatal depression has been associated with adverse birth and child development outcomes. DNA methylation (DNAm) may aid in understanding this association. In this project, we analyzed associations between prenatal depression and DNAm from cord blood from participants of the South African Drakenstein Child Health Study. We examined DNAm in an epigenome wide association study (EWAS) of 248 mother child pairs. DNAm was measured using the Infinium MethylationEPIC (N=145) and the Infinium HumanMethylation450 (N=103) arrays. Prenatal depression scores, obtained with the Edinburgh Postnatal Depression Scale (EPDS) and the Beck Depression Inventory II (BDI-II), were analyzed as continuous and dichotomized variables. We used linear robust models to estimate associations between depression and newborn DNAm, adjusted for measured (smoking status, household income, sex, preterm birth, cell type proportions, and genetic principal components) and unmeasured confounding using Cate and Bacon algorithms. Bonferroni correction was used to adjust for multiple testing. DMRcate was used to test for differentially methylated regions (DMRs). Differential DNAm in *GNAS* (cg22798925, Δ beta per IQR(EPDS)=0.0066, p= 1.06 × 10^−7^) was significantly associated with EPDS. For dichotomized BDI-II thresholds, Differential DNAm in *CTNNA2* (cg04859497, Δ beta=-0.064, p= 8.09 × 10^−10^) and *OSBPL10* (cg27278221, Δ beta=-0.020, p= 5.40 × 10^−8^) was significantly associated with the dichotomized BDI-II variables. Eight DMRs were associated with at least two depression scales. Further studies are needed to replicate these findings and investigate their biological impact.

## Introduction

Prenatal depression affects about 15% to 65% of women around the world with a higher percentage in low to middle income countries (LMICs) than high income countries (HICs)^1^. Adverse birth and child development outcomes, such as low birth weight, pre-term birth, and developmental delay, have been observed in children whose mothers experienced prenatal depression^1,2^. Epigenetics has been hypothesized to play a role in this association. Prenatal development is a crucial and vulnerable time for the epigenome due to epigenetic reprogramming that occurs for both DNA methylation (DNAm) and histone modifications during this time^3^. With the exception of imprinted genes, the epigenome is reprogrammed by the global decrease in DNAm pre-implantation and then increase in DNAm following implantation for processes such as organogenesis^4^. Prenatal exposures such as tobacco smoke^5^, maternal stress^6^, or toxins^7^ can affect the child’s epigenome during prenatal development. Changes in the infant’s epigenetic mechanisms, such as DNAm, as a result of prenatal depression may provide insight into this association either as a biomarker or as a possible mediating factor in biological pathways.

Previous studies investigating the association between prenatal depression and differential DNAm have focused on candidate genes such as *NR3C1* and *SLC6A4* ^8-9^. Children exposed to prenatal depression have been shown to have increased DNAm in *NR3C1* and decreased DNAm in *SLC6A4*^8-9^. Epigenome wide association studies (EWAS) have also investigated associations between prenatal depression and differential DNAm^10-11^. Two of these studies found a combined total of 5 CpG sites (cg08667740, cg22868225, cg06808585, cg05245515, cg15264806) and 39 differentially methylated regions (DMRs) associated with prenatal depression^10-11^. These previous studies included mother child pairs only from high income countries including Norway, the Netherlands, the United Kingdom, and the United States^10-12^.

As women from LMICs are particularly vulnerable to prenatal depression^1^, this study aimed to investigate this association using the Drakenstein Child Health Study (DCHS), a population based birth cohort in South Africa^13-14^. This cohort is representative of several aspects of the LMIC context and allows for the study of potential associations between prenatal depression and DNAm in this setting^13^.

## Materials and Methods

### Study population

The study population consisted of 248 mother child pairs from the DHCS cohort with data available for the Edinburgh Postnatal Depression Scale (EPDS) and the Beck Depression Inventory II (BDI-II) scores, cord blood DNAm, and covariates. Participants were recruited between March 2012 and March 2015 from two primary care clinics, TC Newman or Mbekweni^13,15^. Mothers were enrolled during their second trimester and followed until the child was at least five years old^13-16^.

Ethical approval was given from the Human Research Ethics Committee of the Faculty of Health Sciences of University of Cape Town for human subjects’ research and written consent was obtained from the mothers^13,15^.

### DNA methylation measurements

DNA methylation was measured from cord blood collected at delivery by either the MethylationEPIC BeadChips (n=145) or the Illumina Infinium HumanMethylation450 BeadChips (n=103)^13,15^. The subgroup that was selected for the second set of DNAm analyses (EPIC, n=145) was enriched for maternal trauma exposure/post-traumatic stress disorder (PTSD).

Pre-processing and statistics were done using R 3.5.1. Raw iDat files were imported to RStudio where intensity values were converted into beta values. The 450K array had 426,378 probes while the EPIC array contained 781,536 probes. Pre-processing was performed in each array separately but with identical pre-processing steps. Background subtraction, color correction and normalization were performed using the preprocessFunnorm function. After sample and probe filtering, 120 samples and 426,378 probes remained for the 450K dataset with 153 samples and 781,536 probes with the EPIC dataset. Batch effects were removed using ComBat from the R package sva. Cord blood cell type composition was predicted using the most recent cord blood reference data set and the IDOL algorithm and probe selection.

### Depression measurements

Prenatal depression was assessed with both the EPDS and BDI-II administered at 28 to 32 weeks’ gestation^13,15^. The EPDS scale has 10 questions, score ranges from 0 to 30, and was designed to screen for postnatal depression^17^. This scale has been verified for prenatal depression in an African setting^18^. The BDI-II scale has 21 questions, score ranges from 0 to 63, and is used to screen for depression^19^. The BDI-II scale has been validated for prenatal depression^20^ and used for prenatal depression in countries such as Ethiopia and Kenya^21-22^. For EPDS, thresholds of 10 and 13 are commonly used to screen for depression with 10 having a higher sensitivity and 13 having a higher specificity^23^. For BDI-II, the lower threshold of 14 is the threshold for mild depression, and a higher threshold of 20 is the threshold for moderate depression^19^.

### Statistical analysis

The association between prenatal maternal depression and newborn differential methylation at individual CpG sites and DMRs was assessed in epigenome-wide association studies (EWAS). We conducted EWAS for the 450K and EPIC data separately, followed by a meta-analysis to combine the results of the CpG sites that were measured with both arrays. For each of the EWAS analyses, a multivariable robust linear regression model with empirical Bayes using the limma R package was fitted^24^. The dependent variable was cord blood DNAm, with depression variables as the independent variable while adjusting for the following covariates: mother’s smoking status, household income, sex of the child, preterm birth (<37 weeks), first three cell type principal components (PCs) which explained 90% of heterogeneity due to cell type^25^, and first five genotype PCs for population stratification. A sensitivity analysis was performed to determine the effect of including HIV exposure as potential confounder in the model. P-values were additionally adjusted for bias and unmeasured confounding using the Bacon and Cate R packages respectively^26^. The continuous depression scale was used as primary outcomes, followed by analyses of the dichotomized variables (screening for depression) as secondary outcomes. To account for multiple testing, the Bonferroni threshold was used for statistical significance (EPIC: 0.05 / 781536 CpGs = 6.40 × 10^−8^, 450K: 0.05 / 426378 CpGs =1.17 × 10^−7^, meta-analysis: 0.05 / 386685 CpGs = 1.29 × 10^−7^). Fine-mapping of our epigenome-wide associations was done with the R package comet, which displays the region surrounding any significant CpG sites^27^. DMRs were assessed from the meta-analysis of the overlapping CpG sites from EPIC and 450K using the R package DMRcate (version 1.20.0)^28^. The input files for these analyses included the regression coefficients, standard deviations, and p-values from the single-CpG analyses. DMRs were defined by requiring at least two CpG sites within 1,000 bps apart and the region having an FDR correct p-value <0.01. Furthermore, we used robustness of DMRs across different depression scales as an additional validation criterion.

For any significant CpG sites or DMRs, we looked up the correlations between blood and brain DNA methylation using the public data source IMAGE-CpG, which is based on blood, saliva, buccal, and live brain tissue samples from 27 patients with medically intractable epilepsy undergoing brain resection^29^.

## Results

### Study population characteristics

The analysis sample included 248 mother child pairs with complete information for depression scores, cord blood DNAm, and relevant covariates (Table 1). DNAm was measured in cord blood of 145 infants (58%) using the EPIC array and in 103 infants (42%) using the 450K array. Overall, 44% of the infants were female and around 11% of births were born preterm (before 37 weeks). About 21% of the mothers were smokers during pregnancy with a higher proportion of smokers in 450K data than in the EPIC data. The average EPDS score was 10.52 (sd = 5.09) with 56% defined as depressed according to the threshold of 10 and 31% according to the threshold of 13. The average BDI-II score among mothers was 13.19 (sd = 11.17) with 44% and 25% defined as depressed according to the thresholds of 14 or 20, respectively. The women in the 450K array group (n=103) tended to have higher depression scores (EPDS and BDI-II) than in the EPIC data.

**Table 1.** Population characteristics for the total population and stratified by arrays

| Characteristic | Combined (n=248) | EPIC (n=145) | 450K (n=103) |
| --- | --- | --- | --- |
| **Female, n (%)** | 110 (44.35%) | 67 (46.21%) | 43 (41.75%) |
| **Preterm birth (<37 weeks), n (%)** | 27 (10.89%) | 15 (10.34%) | 12 (11.65%) |
| **Household income** |  |  |  |
| <R1,000/month, n (%) | 98 (39.52%) | 58 (40.00%) | 40 (38.83%) |
| R1,000-R5,000/month, n (%) | 107 (43.15%) | 64 (44.83%) | 43 (41.75%) |
| >R5,000/month, n (%) | 43 (17.33%) | 23 (15.17%) | 20 (19.42%) |
| **Maternal smoking, n (%)** | 53 (21.37%) | 27 (18.62%) | 27 (26.21%) |
| **EPIC Array, n (%)** | 145 (58.47%) | 145 (100.00%) | 0 (0.00%) |
| **EPDS Continuous, mean (sd)** | 10.52 (5.09) | 10.30 (4.64) | 10.83 (5.68) |
| **EPDS Threshold 10, n (%)** | 140 (56.45%) | 83 (57.24%) | 58 (56.31%) |
| **EPDS Threshold 13, n (%)** | 78 (31.45%) | 40 (27.59%) | 38 (36.89%) |
| **BDI-II Continuous, mean (sd)** | 13.19 (11.17) | 11.26 (10.53) | 15.89 (11.55) |
| **BDI-II Threshold 14, n (%)** | 108 (43.55%) | 54 (37.24%) | 54 (52.43%) |
| **BDI-II Threshold 20, n (%)** | 62 (25.00%) | 26 (17.93%) | 36 (34.95%) |

### Maternal Depression Scores and Newborn DNAm

After accounting for bias and measured and unmeasured confounding, the EWAS of the individual 450K and EPIC data for continuous depression variables (EPDS and BDI-II scores) did not result in any significant CpG sites (Figures S1-S4). After combining the overlapping CpG sites from both arrays in a meta-analysis, we found significant associations between differential DNAm in cg22798925 (*GNAS)* and the EPDS score (Δ beta per IQR of EPDS = 0.0066, p-value = 1.06 × 10^−7^) (Figure 1). This CpG site had suggestive p-values for the continuous BDI (Δ beta per BDI-II total IQR = 0.0056, p-value = 1.27 × 10^−4^) and both threshold EPDS variables (threshold-10: Δ beta = 0.0109, p-value = 1.52 × 10^−7^, threshold-13: Δ beta = 0.0088, p-value = 1.47 × 10^−4^) and nominally significant p-values for the threshold BDI variables (Table 2). Associations with DNAm in cg22798925 were similar for data from both arrays (Figure 2). For CpG sites that reached p-values less than 5 × 10^−4^ for at least one of the depression scales in the meta-analysis, beta estimates for the BDI-II and EPDS continuous variables were correlated (Figure S5).

**Table 2.**
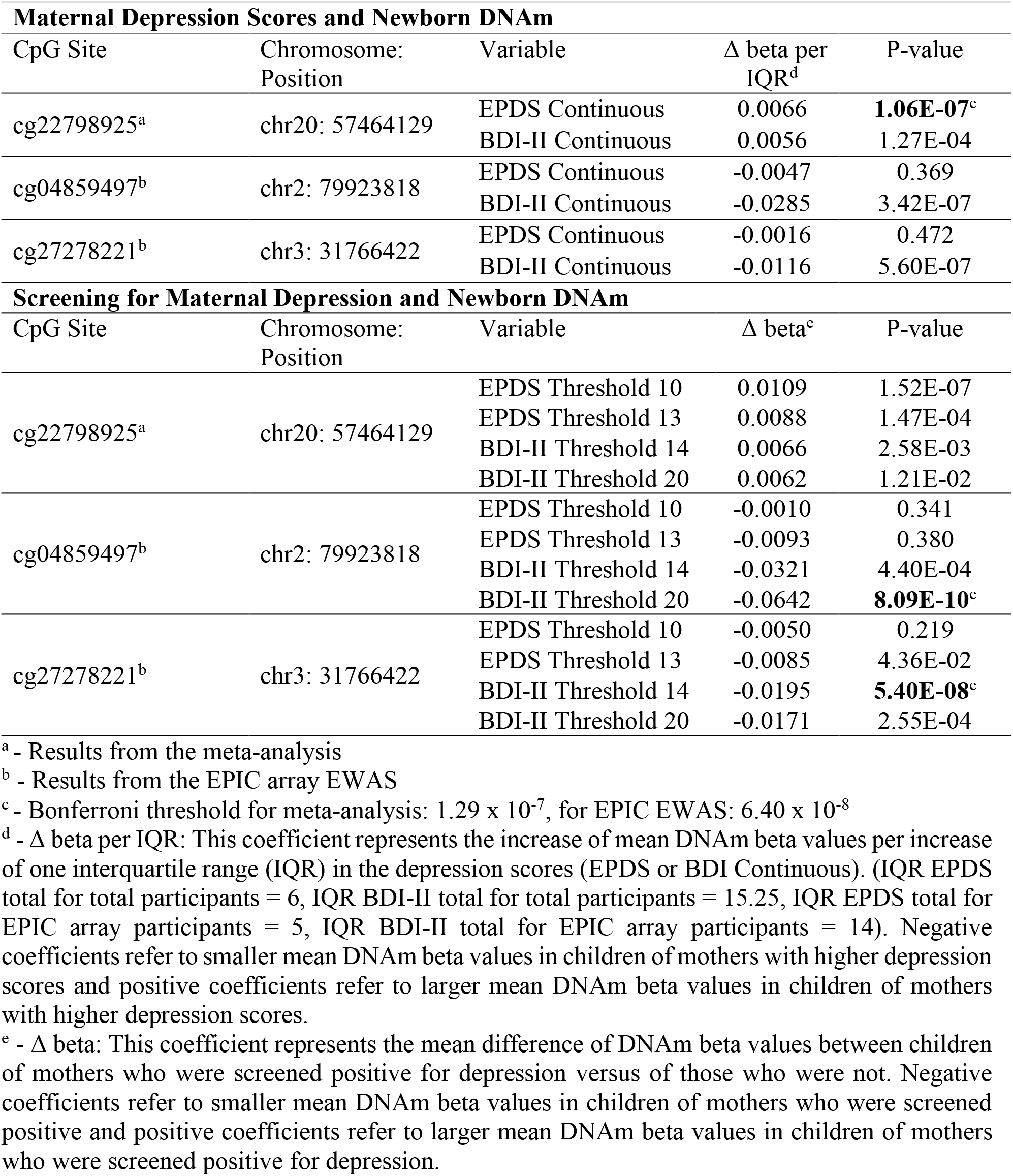
Effect sizes and p-values of significant CpG sites for each depression variable

**Figure 1.**
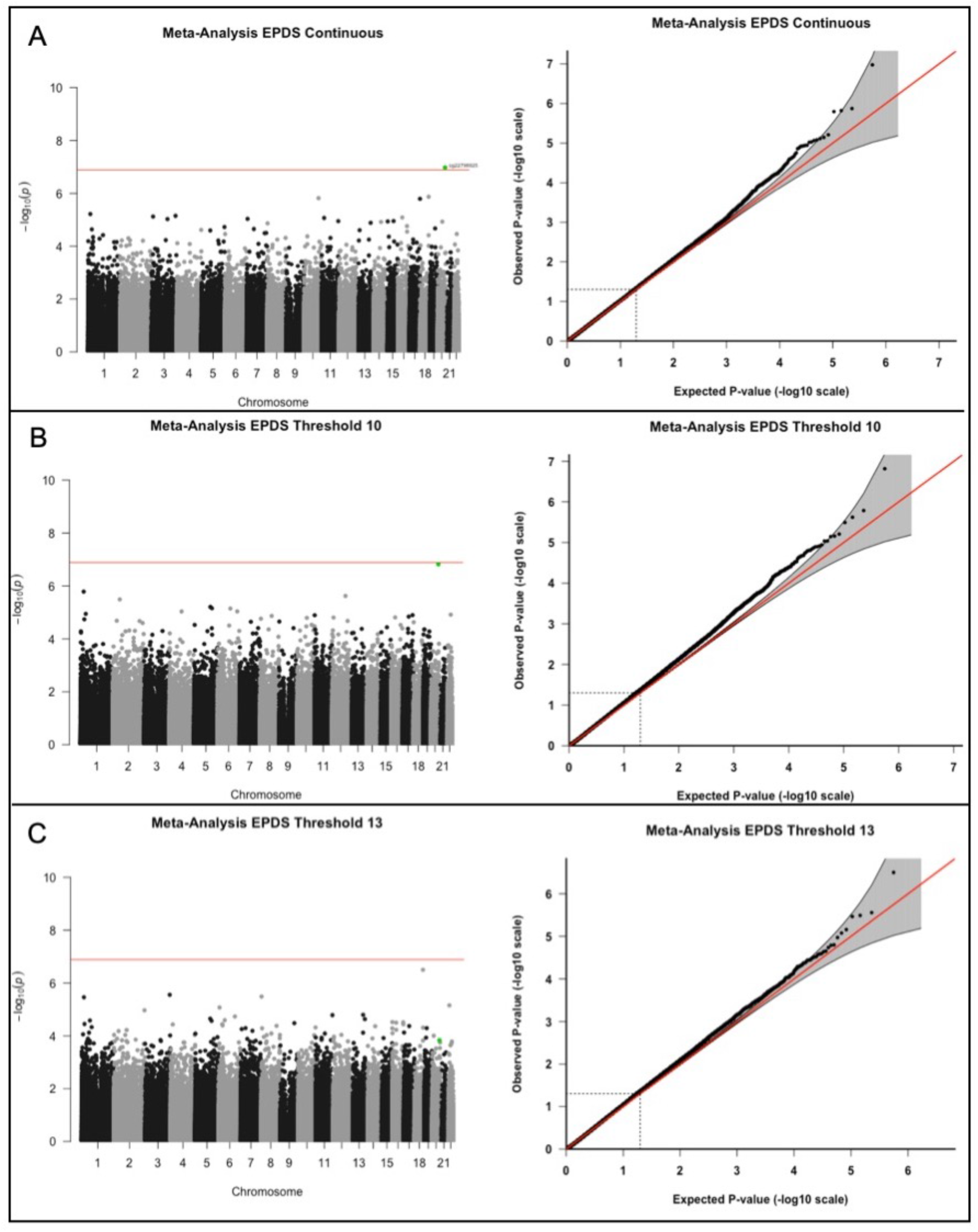
Manhattan and QQ-plots for significant CpG sites with cg22798925 highlighted. Adjusted for covariates: mother’s smoking status, average household income, child’s sex, preterm birth (<37 weeks), first three cell type PCs, and first five genotype PCs. Unmeasured confounding and bias were adjusted with Cate and Bacon R packages. Bonferroni threshold = 1.29 × 10^−7^. A) Meta-analysis results from the EPDS continuous score. B) Meta-analysis results from the EPDS 10 threshold. C) Meta-analysis EWAS results from the EPDS 13 threshold.

**Figure 2.**
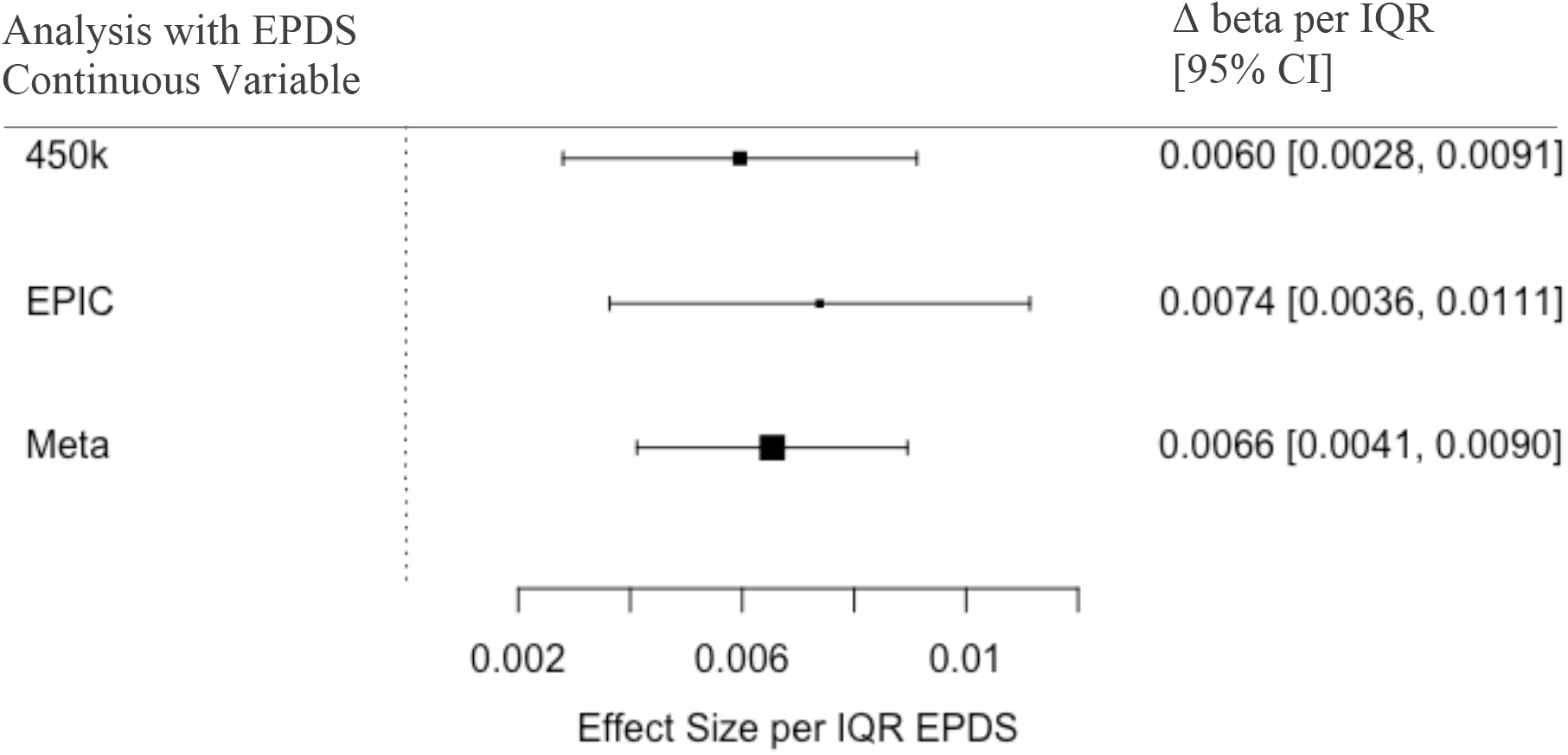
Forest plot indicating effect sizes per IQR of the EPDS scores and 95% confidence intervals per interquartile range (IQR) (IQR EPDS total = 6, IQR BDI-II total = 15.25) for cg22798925 for both EWAS analyzes (450K alone and EPIC alone) and the meta-analysis for the EPDS continuous variable. Adjusted for covariates: mother’s smoking status, average household income, child’s sex, preterm birth (<37 weeks), first three cell type PCs, and first five genotype PCs. Unmeasured confounding and bias were adjusted with Cate and Bacon R packages.

The meta-analysis resulted in 21 DMRs for the EPDS continuous score and 16 DMRs for the BDI-II continuous score (Tables S1-S2). The DMR, chr18: 67069959-67070461, was significant for both depression variables (EPDS: max Δ beta estimate = -0.0026, p-value = 4.41 × 10^−10^, BDI-II: max Δ beta estimate = -0.0011, p-value 3.87 × 10^−6^) (Table 3).

**Table 3:** Max effect sizes and p-values from meta-analysis for DMRs significant in two or more variables

| **Maternal Depression Scores and Newborn DNAm** |  |  |  |  |
| --- | --- | --- | --- | --- |
| DMRs | # CpGs | Variable | Max Effect | P-value* |
| chr18: 67069959-67070461 | 6 | EPDS Continuous | -0.0026 | 4.41E-10 |
|  |  | BDI-II Continuous | -0.0011 | 3.87E-06 |
| chr7: 155174726-155175340 | 4 | BDI-II Continuous | 0.0016 | 3.47E-05 |
| chr6: 33047944-33049360 | 16 | BDI-II Continuous | 0.0021 | 1.79E-11 |
| chr8: 70378380-70378994 | 7 | BDI-II Continuous | 0.0012 | 2.36E-04 |
| chr11: 65190825-65191707 | 4 | BDI-II Continuous | 0.0025 | 2.74E-04 |
| chr12: 104697193-104697983 | 12 | BDI-II Continuous | 0.0011 | 2.70E-04 |
| chr15: 98195808-98196247 | 4 | EPDS Continuous | -0.0032 | 1.11E-05 |
| chr19: 18698825-18699631 | 9 | EPDS Continuous | -0.0034 | 1.46E-05 |
| **Screening for Maternal Depression and Newborn DNAm** |  |  |  |  |
| DMRs | # CpGs | Variable | Max Effect | P-value |
| chr18: 67069959-67070461 | 6 | EPDS Threshold 10 | -0.0253 | 6.10E-04 |
|  |  | EPDS Threshold 13 | -0.0232 | 3.62E-10 |
|  |  | BDI-II Threshold 20 | -0.0241 | 1.86E-07 |
| chr7: 155174726-155175340 | 4 | BDI-II Threshold 14 | 0.0326 | 1.11E-04 |
|  |  | BDI-II Threshold 20 | 0.0378 | 1.98E-04 |
| chr6: 33047944-33049360 | 16 | BDI-II Threshold 20 | 0.0471 | 4.05E-08 |
| chr8: 70378380-70378994 | 7 | BDI-II Threshold 20 | 0.0336 | 1.19E-05 |
| chr11: 65190825-65191707 | 4 | BDI-II Threshold 14 | 0.0640 | 2.93E-06 |
| chr12: 104697193-104697983 | 12 | BDI-II Threshold 14 | 0.0274 | 1.00E-08 |
| chr15: 98195808-98196247 | 4 | EPDS Threshold 10 | -0.0335 | 1.37E-04 |
| chr19: 18698825-18699631 | 9 | EPDS Threshold 10 | -0.0415 | 1.37E-04 |

\*Minimum FDR adjusted p-value from CpGs forming the significant DMR

### Screening for Maternal Depression and Newborn DNAm

The EWAS of the EPIC data resulted in cg04859497 and cg27278221 being statistically significant for the BDI-II threshold 20 (Δ beta = -0.064, p-value = 8.09 × 10^−10^) and BDI-II threshold 14 (Δ beta = -0.020, p-value =5.40 × 10^−8^), respectively (Table 2). These CpG sites are unique to the EPIC array and not available on the 450K array. The p-values for both of the CpG sites were suggestive for the continuous BDI-II depression variables but not for the EPDS depression variables (Table 2). We did not find any significant associations between the dichotomized screening variables for maternal depression and single CpG sites in the 450K analyses nor in the meta-analysis.

The meta-analysis resulted in 67 DMRs for the binary variables (EPDS threshold-10: 14 DMRs, EPDS threshold-13: 19 DMRs, BDI-II threshold-14: 19 DMRS, BDI-II threshold-20: 19 DMRs) (Tables S3-S6). Eight DMRs were significant for more than one variable, continuous or dichotomized, and two of the eight DMRs, chr18: 67069959-67070461 and chr7: 155174726-155175340, were significant for more than one dichotomized variable (Table 3).

## Discussion

In this study of infants from a peri-urban region in a low-resourced community in South Africa, we found prenatal depression to be associated with differential methylation in *GNAS, CTNNA2, OSBPL10*, and within multiple DMRs measured in cord blood of newborns from the Drakenstein Child Health Study.

### Comparison with previous studies

The association between maternal prenatal depression and differences in infant DNAm is not completely understood. Previous studies measuring DNAm from cord blood have shown mixed results. A study by Viuff et al. (2018), which used the EPDS threshold-12 variable to screen for prenatal depression, found differential methylation in two CpG sites to be associated with prenatal depression while a study by Cardenas, A. et al. (2019), which used the Brief Symptom Inventory threshold-0.80 variable to screen for prenatal depression, found differential methylation in three different CpGs to be associated with prenatal depression^10-11^. However, the significant sites for both of these studies were unable to be replicated using the Generation R study^10-11^. The five CpG sites identified in previous studies were not significantly associated with prenatal depression in our cohort, which is in line with the results from the Generation R study (Table S7-S8). As for DMRs, Cardenas et al. (2019) did not find any DMRs significantly associated with prenatal depression^11^. However, Viuff, et al. (2018) found 39 DMRs to be associated with prenatal depression^10^. Of these 39 DMRs, a DMR containing 8 CpGs, chr8:70378380-70378995^10^, overlaps with a 7 CpG DMR, chr8: 70378380-70378994, found to be significantly associated with the BDI-II continuous and BDI-II threshold-20 in our study. This replicated DMR in chr8 was previously found to be significant for mid-pregnancy maternal depression, which is defined as depression between 18 to 32 weeks gestation^10^. This overlaps with the time maternal depression was assessed in our study, which was between 28 to 32 weeks gestation^30^. The DMR in chr8: 70378380-70378994 overlaps with the promoter region for *SULF1* which codes for the extracellular sulfatase Sulf-1 and is involved in regulating heparin sulfate (HS)-dependent signaling pathways^31^. In mice, deficiencies in *SULF1* were associated with impaired neurite outgrowth, providing evidence for the role of *SULF1* in nervous system development^31-32^. One site within this DMR, cg07051728, was found to have a significant correlation between DNAm in brain tissue and DNAm in blood (Table S9).

### Maternal Depression Scores and Newborn DNAm

In our meta-analysis of overlapping CpG sites from the 450K and EPIC arrays, we found a CpG site located within *GNAS*, which has been previously associated with prenatal maternal stress at a different CpG site, to be associated with prenatal depression score EPDS ^33^. This gene is complex, containing four promoters and producing multiple transcripts while also being involved in genomic imprinting^33^. Imprinted genes play an important role and are vulnerable to maternal exposures during prenatal development^33-34^. This CpG site is located in an intron and 1bp upstream the SNP, rs191456206. This SNP has G as the major allele and with C and T as the minor alleles. However, the C and T alleles appear to be rare with a minor allele frequency of 1.6 × 10^−5^ for the C allele and 0.0 for the T allele. This CpG site is located within a promoter regulatory region. *GNAS* is involved in hormone pathways through aiding in the production of cyclic AMP with highest expression in the pituitary and thyroid glands^35^. Defects in maternal imprinting have been found to be associated with pseudohypoparathyroidism type 1b (PHP1B), a disorder with resistance to parathyroid hormone as a result of reduced expression^36^. *GNAS* is also thought to play a role in fetal growth due to certain mutations being associated with severe intrauterine growth retardation (IUGR) and other mutations along with loss of methylation being associated with increased fetal growth^37^. Due to *GNAS’s* role in fetal development and changes in methylation being associated with increased fetal growth, investigating the methylation status of *GNAS* may be helpful for researching biological pathways between prenatal depression and adverse birth and development outcomes in children.

The meta-analysis resulted in 21 DMRs for the continuous EPDS score and 16 DMRs for the continuous BDI-II score with a DMR in chr18: 67069959-67070461 being significantly associated with all depression variables except for the BDI-II threshold-14 variable. This DMR overlaps with the promoter region for docking protein 6 (*DOK6)*, specifically the protein coding transcript DOK6-001, previously shown to perform a role in Ret-mediated neurite growth^38^ and nervous system development through NT-3 mediation in mice^39^. Up to now, there has not been human research for this protein and neurodevelopment. However, in human tissues, *DOK6* has been shown to have high expression in the fetal brain^38^. This DMR contained one site with a significant correlation for DNAm between brain tissue and blood (Table S9). This DMR may be important for studying adverse developmental outcomes in children born to mothers who experienced prenatal depression. Two DMRs, chr6: 33047944-3304960 and chr15: 98195808-98196247 were significantly associated with a continuous and threshold variable in the meta-analysis however no clear link between these sites and adverse birth or developmental outcomes or prenatal depression has been reported in the literature.

### Screening for Maternal Depression and Newborn DNAm

In the EPIC array specific analysis, a single CpG site, cg27278221, was associated with the BDI-II threshold-14 variable and was suggestive with the remaining BDI-II variables. This site is located within a CTCF transcription factor binding site in *OSBPL10*, a gene involved in lipid metabolism^40^. A single CpG site found from the EPIC array specific analysis was significantly associated with the BDI-II threshold-20 variable and also suggestive for the other BDI-II variables. This site is located in the second intron within *CTNNA2* which codes for the catenin alpha-2 protein which plays an important role in neurodevelopment by acting as a regulator for actin branching, with mutations in this gene associated with a neuronal migration disorder^41^. *CTNNA2* has been shown to have higher expression in the brain than most other tissues^41^. However, the CpG site, cg04859497, was not found to have a significant correlation for DNAm across brain tissue and blood (Table S9). As a result, it is unknown whether this site would also have differential methylation in cells within prenatal brain tissue due to prenatal depression.

The meta-analysis resulted in 67 DMRs for the dichotomized variables (EPDS threshold-10: 14 DMRs, EPDS threshold-13: 19 DMRs, BDI-II threshold-14: 19 DMRS, BDI-II threshold-20: 19 DMRs) and eight of these were significant for more than one depression variable (including the replicated DMR in chr8: 70378380-70378994 discussed above). The DMR, chr11: 65190825-65191707 was found to be significant for both BDI-II continuous and BDI-II threshold-14 and overlaps with *NEAT1* which codes for a long non-coding RNA important for forming paraspeckles within cells^42^. Altered expression of *NEAT1* has been associated with cancer, neurodegenerative disorders, psychiatric diseases, and neuronal excitability^42-43^. None of the CpG sites in this DMR, however, had a significant correlation for DNAm across brain tissue and blood (Table S9). Differences in *NEAT1* expression have also been shown to be associated with IUGR^44^. Long non-coding RNA from *NEAT1* was found to be up regulated in the fetal part of the placenta for infants with IUGR^44^. While it does not appear to be known whether DNAm plays a direct role in the increased *NEAT1* expression, differential methylation in *NEAT1* may provide insight between prenatal depression and low birth weight due to the association between *NEAT1* and IUGR^44^. The DMR chr12: 104697193-104697983 was significant for the BDI-II continuous and BDI-II threshold-14 variables and overlaps with the *TXNRD1* and *EID3* genes. In mice, evidence has been provided for a possible role of *TXNRD1*, which codes for cytosolic thioredoxin reductase, in brain development^45^. *EID3* codes for E1A-like inhibitor of differentiation 3 (EID3) which has been shown to inhibit CBP transcriptional activity^46^ and has also been thought to aid in the regulation of DNMT3A to affect the methylation status in umbilical cord mesenchymal stem cells while the cells transdifferentiate to neural stem-like cells^47^. The DMR, chr7: 155174726-155175340, and chr19: 18698825-18699631 were significant for more than one depression variable, however, the connection between these regions and adverse outcomes due to prenatal depression is not clear. Overall, these sites and DMRs may be useful for investigating the biological pathway for the association between maternal prenatal depression and adverse birth and child development outcomes.

### Strengths and Limitations

Our study has several strengths. Previous studies have only focused on one dichotomized depression scale^10-11^. In our study, we used more than one scale with continuous and dichotomized variables to reflect the complexity of depression and to validate the robustness of our findings across different depression scales. Another strength lies in the study population being of African and mixed ancestry and from a low to middle income country, underrepresented populations among genetic and epigenetic studies^48-49^. A major contributor to the variation in DNA methylation is genetic variation. This study includes genome-wide genotype data which was used to correct for population stratification. Another issue which plagues EWASs is unknown confounding. This study used Cate and Bacon to control for bias and unmeasured confounding, which are state-of-the-art confounder adjustment methods based on the calculation of surrogate variables and the empirical null distribution, respectively^27^.

This study does include limitations such as a relatively small study size which reduces the power to detect differences in methylation^50^. The depression scores were collected at a single time point resulting in the inability to know the complete timeframe for the onset and duration of the prenatal depression^30^. Also, this analysis is based on depression scales instead of clinical diagnosis of prenatal depression, though the depression scales are used to screen for probable depression but are not equivalent to clinical diagnosis^21^. Fourthly, having these two datasets measured on different array platforms resulted in EPIC specific sites not being assessed in the meta-analysis excluding these results from benefiting of larger samples sizes. Another limitation in this study is the use of a heterogenous tissue. Although variance in the proportions of cell types were controlled for using methylation predicted estimated cell counts^51^, the specific cell type origin for a given change in methylation was impossible to determine. This is an issue which is alleviated using techniques such as single cell methylation measurements. A large limitation in most EWASs is in interpreting what changes in methylation actually mean. Even though there have been studies showing the impact of DNAm on gene expression, interpretations must be taken with caution. Additionally, many of the genes which were differentially methylated in association with maternal depression in our study have been linked to various neurological outcomes. We know that DNAm varies most greatly between tissues, as establishing cellular identity is one of the main functions of DNAm. As such, methylation status observed in blood cannot be extrapolated to the methylation states in the brain. While there are datasets with matched brain and blood methylation available to help support the link between blood methylation and neurological outcomes, interpretations of differential methylation need to be taken with caution.

## Conclusion

Maternal depression was associated with differential DNAm in *GNAS, CTNNA2, OSBPL10*, and within multiple DMRs. The DMR chr8: 70378380-70378994 has been associated with maternal depression in a previous study^10^. The remaining sites and DMRs, to our knowledge, have not been previously associated with maternal depression. Further research is needed to replicate this finding and to investigate its impact on birth outcomes and child development.

## Data Availability

Data are available upon request (PI: Prof. Heather Zar)

## Acknowledgments

The authors thank the study and clinical staff at Paarl Hospital, Mbekweni and TC Newman clinics, as well as the CEO of Paarl Hospital, and the Western Cape Health Department for their support of the study. The authors thank the families and children who participated in this study.

## Web Resources

Ensembl, NCBI’s Gene resources, Human Protein Atlas (all accessed on 4/9/2021):

- Overlapping genes for DMR chr8: 70378380-70378995: https://grch37.ensembl.org/Homo_sapiens/Location/View?r=8%3A70378380-70378995
- Position in *GNAS* for cg22798925: http://grch37.ensembl.org/Homo_sapiens/Gene/Summary?db=core;g=ENSG00000087460;r=20:57414773-57486247; SNP next to cg22798925: https://www.ncbi.nlm.nih.gov/snp/?term=rs191456206
- rs191456206 minor allele frequency information: https://www.ncbi.nlm.nih.gov/snp/rs191456206#frequency_tab
- *GNAS* expression levels: https://www.proteinatlas.org/ENSG00000087460-GNAS#gene_information
- Overlapping genes for DMR chr18: 67069959-67070461: https://grch37.ensembl.org/Homo_sapiens/Location/View?r=18%3A67069959-67070461
- Overlapping genes for DMR chr6: 33047944-3304960:http://grch37.ensembl.org/Homo_sapiens/Location/Overview?r=6%3A3304960
- -33047944
- Gene classification for *RPL32P1*: http://grch37.ensembl.org/Homo_sapiens/Gene/Summary?db=core;g=ENSG00000224796;r=6:33047228-33047637;t=ENST00000439737
- Protein coded by *HLA-DPa1*: https://www.uniprot.org/uniprot/P20036
- Protein coded by *HLA-DPb1*: https://www.uniprot.org/uniprot/P04440
- Overlapping genes for DMR chr15: 98195808-98196247: http://grch37.ensembl.org/Homo_sapiens/Location/View?r=15:98196029-98196118;db=core
- *OSBPL10* position of cg27278221: http://grch37.ensembl.org/Homo_sapiens/Location/View?r=3%3A31765422-31767422
- *CTNNA2* position of cg04859497: https://grch37.ensembl.org/Homo_sapiens/Location/View?db=core;g=ENSG00000066032;r=2:79900309-79939089;t=ENST00000496558
- Overlapping genes for DMR chr7: 55174726-155175340: http://grch37.ensembl.org/Homo_sapiens/Location/View?r=7%3A155174726-155175340
- Overlapping genes for DMR chr11: 65190825-65191707: http://grch37.ensembl.org/Homo_sapiens/Location/View?r=11%3A65190825-65191707
- Overlapping genes for DMR chr12: 104697193-104697983: http://grch37.ensembl.org/Homo_sapiens/Location/View?r=12%3A104697193-104697983
- Overlapping genes for DMR chr19: 18698825-18699631: http://grch37.ensembl.org/Homo_sapiens/Location/View?r=19%3A18698825-18699631

## Notes

### Competing Interest Statement

The authors have declared no competing interest.

### Funding Statement

The Drakenstein Child Health Study was funded by the Bill & Melinda Gates Foundation (OPP 1017641), Discovery Foundation, Medical Research Council South Africa, National Research Foundation South Africa, CIDRI Clinical Fellowship and Wellcome Trust (204755/2/16/z). Additional support for the DNA methylation work was by the Eunice Kennedy Shriver National Institute of Child Health and Human Development of the National Institutes of Health (NICHD) under Award Number R21HD085849, and the Fogarty International Center (FIC). The content is solely the responsibility of the authors and does not necessarily represent the official views of the National Institutes of Health. Anke Huels is supported the HERCULES Center (NIEHS P30ES019776). Michael Epstein was supported by NIH grant R01 GM117946. Dan Stein and Heather Zar are supported by the South African Medical Research Council (SAMRC). The funders had no role in the study design, data collection and analysis, decision to publish, or preparation of manuscript.

### Author Declarations

Ethical approval was given from the Human Research Ethics Committee of the Faculty of Health Sciences of University of Cape Town for human subjects research and written consent was obtained from the mothers.

## References

1 Dadi AF, Miller ER, Bisetegn TA, Mwanri L. Global burden of antenatal depression and its association with adverse birth outcomes: An umbrella review. BMC Public Health. https://doi.org/10.1186/s12889-020-8293-9 (2020).

2 Deave T, Heron J, Evans J, Emond A. The impact of maternal depression in pregnancy on early child development. BJOG. 115, 1043–51 (2008).

3 Kundakovic M, Jaric I. The epigenetic link between prenatal adverse environments and neurodevelopmental disorders. Genes (Basel). 18, 104 (2017).

4 Thorsell A, Nätt D. Maternal stress and diet may influence affective behavior and stress-response in offspring via epigenetic regulation of central peptidergic function. Environ Epigenetics. 2, dvw012 (2016).

5 Suter MA, Aagaard K. What changes in DNA methylation take place in individuals exposed to maternal smoking in utero? Epigenomics. 4, 115–118 (2012).

6 Kertes DA. et al.. Prenatal Maternal Stress Predicts Methylation of Genes Regulating the Hypothalamic-Pituitary-Adrenocortical System in Mothers and Newborns in the Democratic Republic of Congo. Child Dev. 87, 61–72 (2016).

7 Appleton AA, Jackson BP, Karagas M, Marsit CJ. Prenatal exposure to neurotoxic metals is associated with increased placental glucocorticoid receptor DNA methylation. Epigenetics. 12, 607–615 (2017).

8 Devlin AM, Brain U, Austin J, Oberlander TF. Prenatal exposure to maternal depressed mood and the MTHFR C677T variant affect SLC6A4 methylation in infants at birth. PLoS One. 5, e12201 (2010).

9 Oberlander TF. et al.. Prenatal exposure to maternal depression, neonatal methylation of human glucocorticoid receptor gene (NR3C1) and infant cortisol stress responses. Epigenetics 3, 97–106 (2008).

10 Viuff AC. et al.. Maternal depression during pregnancy and cord blood DNA methylation: findings from the Avon Longitudinal Study of Parents and Children. Transl Psychiatry. 8, 244 (2018).

11 Cardenas A. et al.. Prenatal maternal antidepressants, anxiety, and depression and offspring DNA methylation: Epigenome-wide associations at birth and persistence into early childhood. Clin Epigenetics. 11, 56 (2019).

12 Wikenius E. et al.. Prenatal maternal depressive symptoms and infant DNA methylation: a longitudinal epigenome-wide study. Nord J Psychiatry. 73, 257–263 (2019).

13 Zar HJ, Barnett W, Myer L, Stein DJ, Nicol MP. Investigating the early-life determinants of illness in Africa: the Drakenstein Child Health Study. Thorax. 70, 592–4 (2015).

14 Brittain K, et al. Risk Factors for Antenatal Depression and Associations with Infant Birth Outcomes: Results From a South African Birth Cohort Study. Paediatr Perinat Epidemiol. 29, 505–14 (2015).

15 Stein DJ, et al. Investigating the psychosocial determinants of child health in Africa: The Drakenstein Child Health Study. J Neurosci Methods. 252, 27–35 (2015).

16 Donald K, et al. Drakenstein Child Health Study (DCHS): investigating determinants of early child development and cognition. BMJ Paediatr Open. 2, e000282 (2018).

17 Gibson J, McKenzie-Mcharg K, Shakespeare J, Price J, Gray R. A systematic review of studies validating the Edinburgh Postnatal Depression Scale in antepartum and postpartum women. Acta Psychiatr Scand. 119, 350–64 (2009).

18 Heyningen T, Honikman S, Tomlinson M, Field S, Myer L. Comparison of mental health screening tools for detecting antenatal depression and anxiety disorders in South African women. PLoS One. 13, e0193697 (2018).

19 Graham RM. et al.. Maternal anxiety and depression during late pregnancy and newborn brain white matter development. AJNR Am J Neuroradiol. 41, 1908–1915 (2020).

20 Naja S. et al.. Psychometric properties of the Arabic version of EPDS and BDI-II as a screening tool for antenatal depression: Evidence from Qatar. BMJ Open. 9, e030365 (2019).

21 Alenko A, Dejene S, Girma S. Sociodemographic and Obstetric Determinants of Antenatal Depression in Jimma Medical Center, Southwest Ethiopia: Facility Based Case-Control Study. Int J Womens Health. 12, 557–565 (2020).

22 Opiyo R. et al.. Effect of fish oil omega-3 fatty acids on reduction of depressive symptoms among HIV-seropositive pregnant women: a randomized, double-blind controlled trial. Ann Gen Psychiatry. 17, 49 (2018).

23 Khanlari S, Barnett Am B, Ogbo FA, Eastwood J. Re-examination of perinatal mental health policy frameworks for women signalling distress on the Edinburgh Postnatal Depression Scale (EPDS) completed during their antenatal booking-in consultation: A call for population health intervention. BMC Pregnancy Childbirth. 19, 221 (2019).

24 Ritchie ME. et al.. Limma powers differential expression analyses for RNA-sequencing and microarray studies. Nucleic Acids Res. 34, e47 (2015).

25 Pawlowsky-Glahn V, Egozcue JJ. Compositional data and their analysis: An introduction. Geol Soc Spec Publ. 246, 1–10 (2006).

26 Van Iterson M, Van Zwet E, BIOS Consortium, Heijmans B. Controlling bias and inflation in epigenome- and transcriptome-wide association studies using the empirical null distribution. Genome Biol. 18, 19 (2017).

27 Martin TC, Yet I, Tsai PC, Bell JT. coMET: Visualisation of regional epigenome-wide association scan results and DNA co-methylation patterns. BMC Bioinformatics. 16, 131 (2015).

28 Peters TJ. et al.. De novo identification of differentially methylated regions in the human genome. Epigenetics Chromatin. 8, 6 (2015).

29 Braun P, et al. Genome-wide DNA methylation comparison between human brain and peripheral tissues within individuals. Transl Psychiatry. 9, 47 (2019).

30 Koen N, et al. Psychological trauma and posttraumatic stress disorder: risk factors and associatiosn with birth outcomes in the Drakenstein Child Health Study. Eur J Psychotraumatol. 7, 28720 (2016).

31 Kalus I, et al. Differential involvement of the extracellular 6-O-endosulfatases Sulf1 and Sulf2 in brain development and neuronal and behavioural plasticity. J Cell Mol Med. 13, 4505–21 (2009).

32 Kalus I, et al. Sulf1 and Sulf2 Differentially Modulate Heparan Sulfate Proteoglycan Sulfation during Postnatal Cerebellum Development: Evidence for Neuroprotective and Neurite Outgrowth Promoting Functions. PLOS One. 10, e0139853 (2015).

33 Vangeel EB. et al.. DNA methylation in imprinted genes IGF2 and GNASXL is associated with prenatal maternal stress. Genes Brain Behav. 14, 573–82 (2015).

34 Moore GE. et al.. The role and interaction of imprinted genes in human fetal growth. Philos Trans R Soc Lond B Biol Sci. https://doi.org/10.1098/rstb.2014.0074 (2015).

35 Long X dan, Xiong J, Mo Z hui, Dong C sheng, Jin P. Identification of a novel GNAS mutation in a case of pseudohypoparathyroidism type 1A with normocalcemia. BMC Med Genet. 19, 132 (2018).

36 Poradosu S, Bravenboer B, Takatani R, Jüppner H. Pseudohypoparathyroidism type 1B caused by methylation changes at the GNAS complex locus. BMJ Case Rep. 2016, bcr2016214673 (2016).

37 Bréhin AC. et al.. Loss of methylation at GNAS exon A/B is associated with increased intrauterine growth. J Clin Endocrinol Metab. 100, E623–31 (2015).

38 Crowder R, Enomoto H, Yang M, Johnson E, Milbrandt J. Dok-6, a Novel p62 Dok family member, promotes Ret-mediated neurite outgrowth. J Biol Chem. 279, 42072–81 (2004).

39 Li W. et al.. Downstream of tyrosine kinase/docking protein 6, as a novel substrate of tropomyosin-related kinase C receptor, is involved in neurotrophin 3-mediated neurite outgrowth in mouse cortex neurons. BMC Biol. 8, 86 (2010).

40 Perttila J. et al. OSBPL10, a novel candidate gene for high triglyceride trait in dyslipidemic Finnish subjects, regulates cellular lipid metabolism. J Mol Med (Berl). 87, 825–35 (2009).

41 Schaffer A. et al.. Biallelic loss of human CTNNA2, encoding αN-catenin, leads to ARP2/3 complex overactivity and disordered cortical neuronal migration. Nat Genet. 50, 1093–1101 (2018).

42 Kukharsky M. et al.. Long non-coding RNA Neat1 regulates adaptive behavioural response to stress in mice. Transl Psychiatry. https://doi.org/10.1038/s41398-020-0854-2 (2020).

43 Dong P. et al.. Long Non-coding RNA NEAT1: A Novel Target for Diagnosis and Therapy in Human Tumors. Front Genet. 9, 471 (2018).

44 Gremlich S, et al. The long non-coding RNA NEAT1 is increased in IUGR placentas, leading to potential new hypotheses of IUGR origin/development. Placenta. 35, 44–9 (2014).

45 Soerensen J. et al.. The Role of Thioredoxin Reductases in Brain Development. PLoS One. 3, e1813.(2008)

46 Bavner A, Matthews J, Sanyal S, Gustafsson J, Treuter E. EID3 is a novel EID family member and an inhibitor of CBP-dependent co-activation. Nucleic Acids Res. 33, 3561-9 (2005)

47 Luo L, Chen W, Yin J, Xu R. EID3 directly associates with DNMT3A during transdifferentiation of human umbilical cord mesenchymal stem cells to NPC-like cells. Sci Rep. 7, 40463 (2017).

48 Popejoy A. Genomics is failing on diversity. Nature. doi: 10.1038/538161a (2016).

49 Cronjé H, Elliott H, Nienaber-Rousseau C, Pieters M. Replication and expansion of epigenome-wide association literature in a black South African population. Clin Epigenetics. 12, 6 (2020).

50 Mansell G, et al. Guidance for DNA methylation studies: statistical insights from the Illumina EPIC array. BMC Genomics. 20, 366 (2019).

51 Kaushal A. et al.. Comparison of different cell type correction methods for genome-scale epigenetics studies. BMC Bioinformatics. https://doi.org/10.1186/s12859-017-1611-2 (2017).

