## Supplement for "Association between Maternal Depression during Pregnancy and Newborn DNA Methylation"

**Supplementary Figures**

A	B

C	D

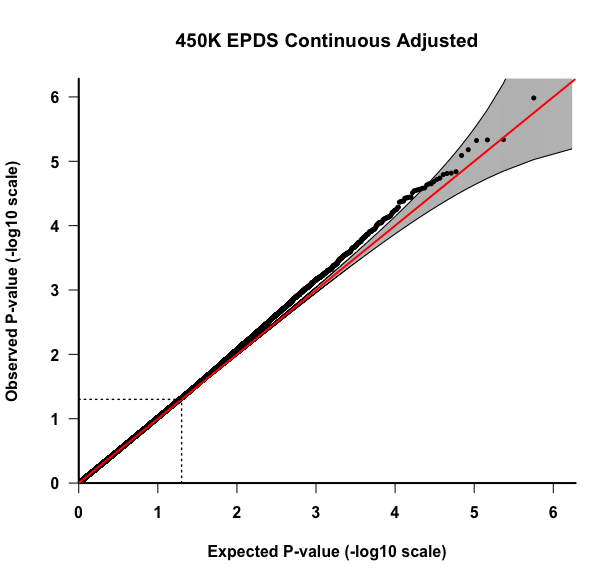

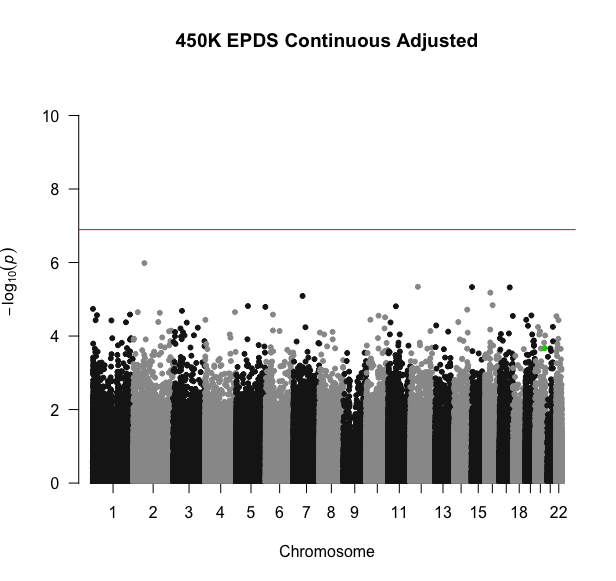

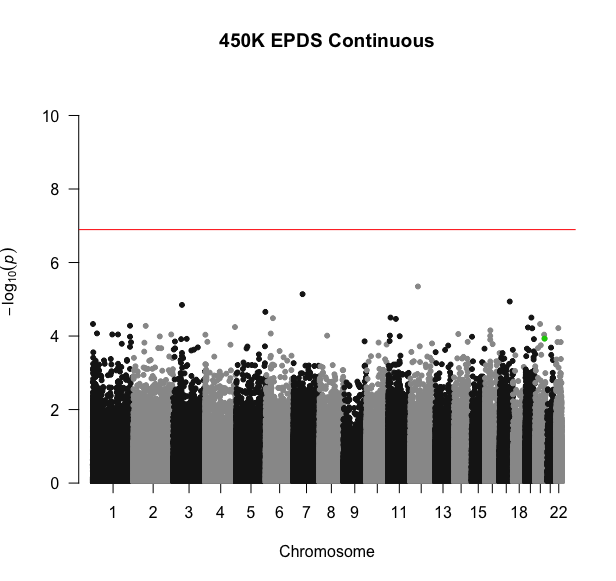

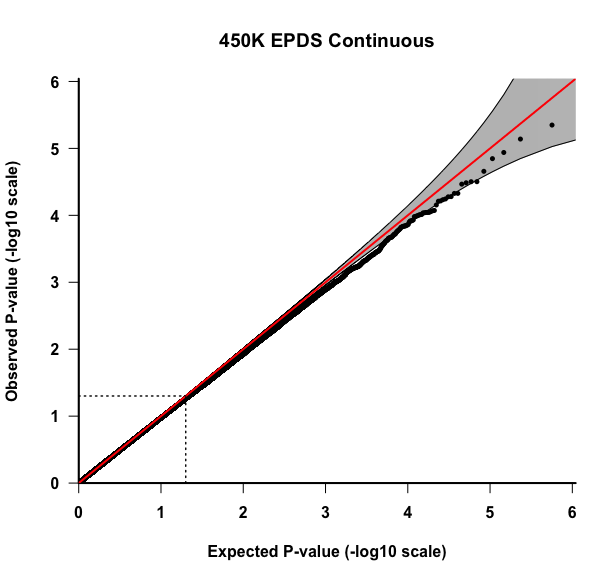

**Figure S1: Results for the 450K EWAS for the EPDS continuous variable.** All association models were adjusted for covariates: mother’s smoking status, average household income, child’s sex, preterm birth (<37 weeks), first three cell type PCs, and first five genotype PCs. A) Plot A is the QQ-plot for the unadjusted p-values. B) Plot B is the Manhattan plot for the unadjusted p-values. The highlighted site is cg22798925. C) Plot C is the QQ-plot for the adjusted p-values using Bacon and Cate. D) Plot D is the Manhattan plot for adjusted p-values using Bacon and Cate. The highlighted site is cg22798925.

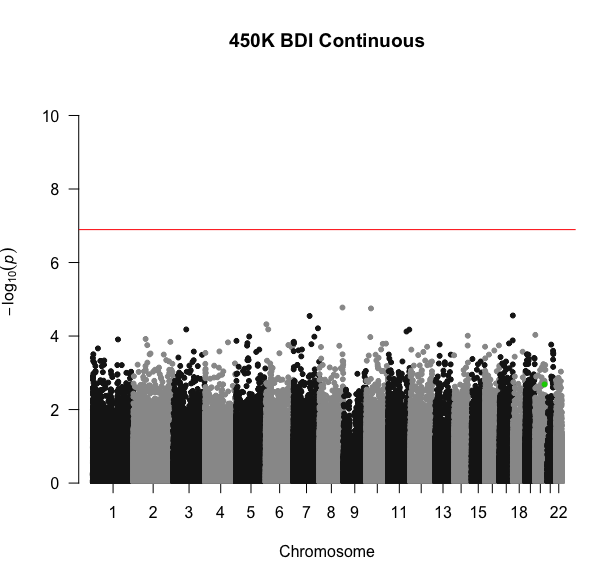

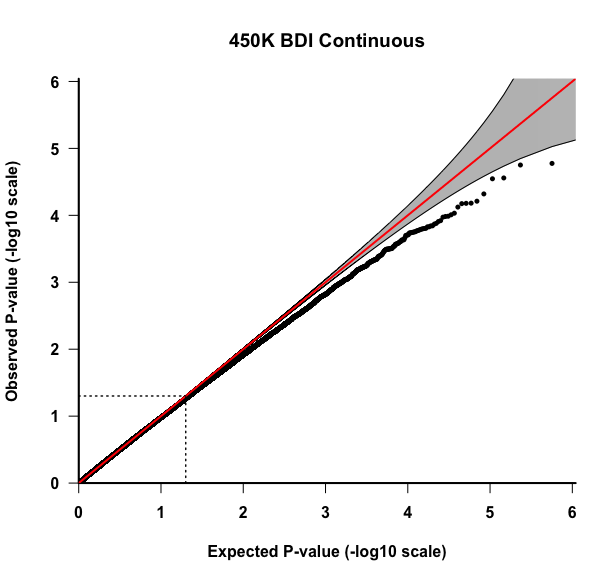

A	B

C	D

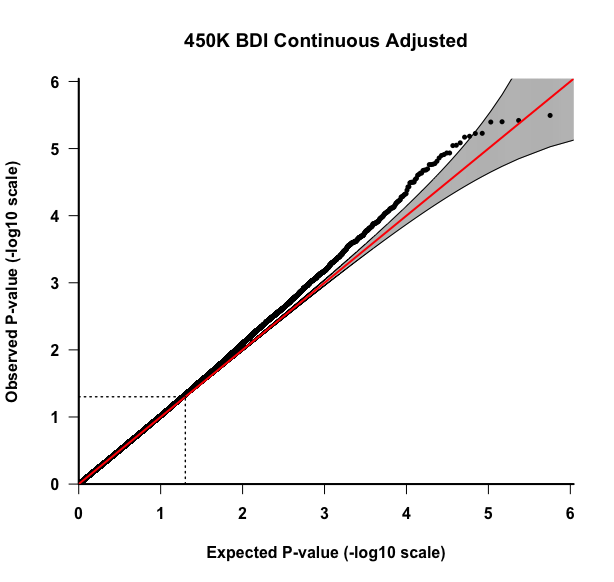

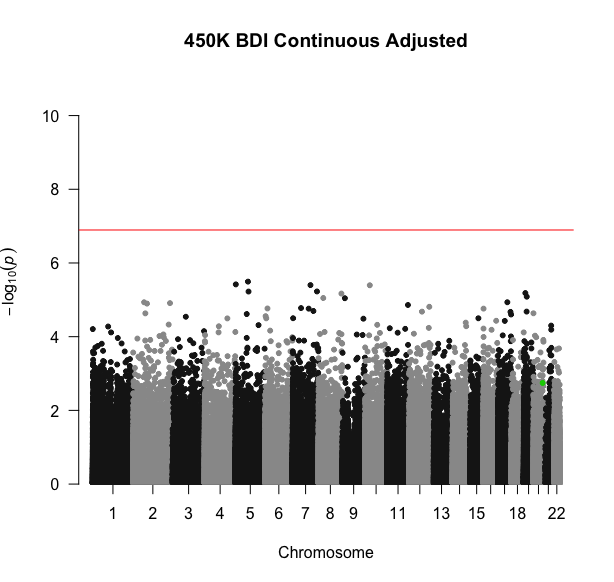

**Figure S2: Results for the 450K EWAS for the BDI-II continuous variable.** All association models were adjusted for covariates: mother’s smoking status, average household income, child’s sex, preterm birth (<37 weeks), first three cell type PCs, and first five genotype PCs.. A) Plot A is the QQ-plot for the unadjusted p-values. B) Plot B is the Manhattan plot for the unadjusted p-values. The highlighted site is cg22798925. C) Plot C is the QQ-plot for the adjusted p-values using Bacon and Cate. D) Plot D is the Manhattan plot for adjusted p-values using Bacon and Cate. The highlighted site is cg22798925.

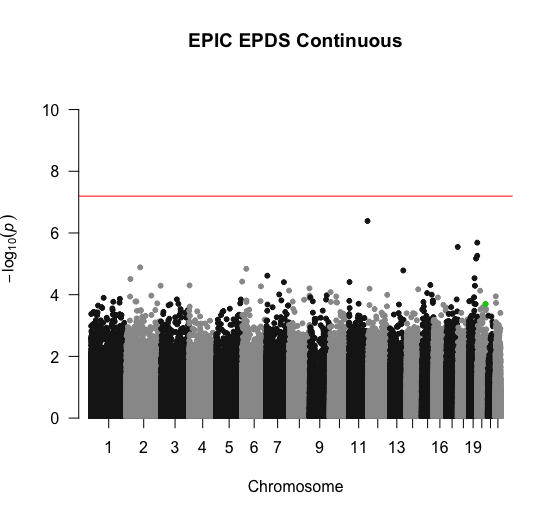

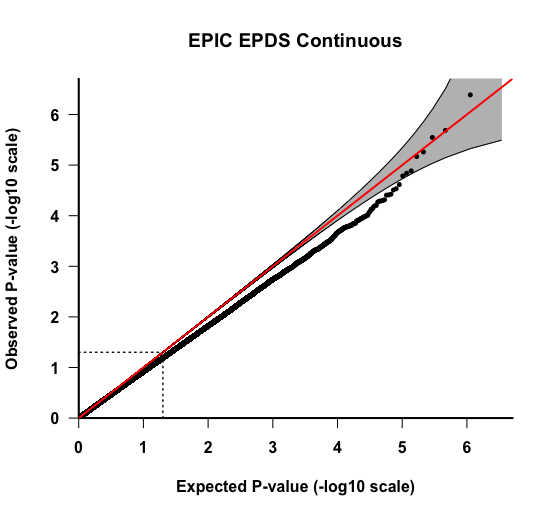

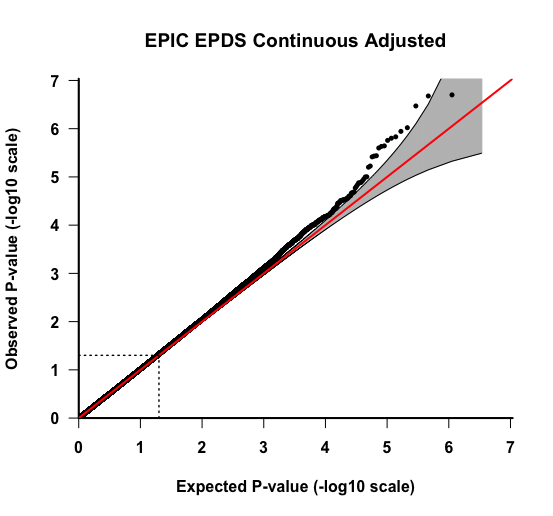

A	B

C	D

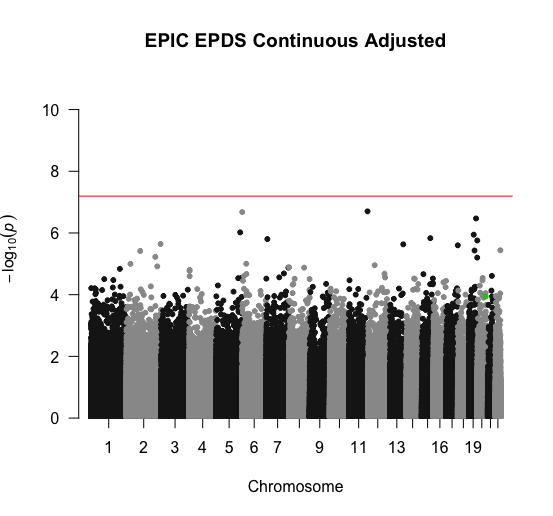

**Figure S3: Results for the EPIC EWAS for the EDPS continuous variable.** All association models were adjusted for covariates: mother’s smoking status, average household income, child’s sex, preterm birth (<37 weeks), first three cell type PCs, and first five genotype PCs.. A) Plot A is the QQ-plot for the unadjusted p-values. B) Plot B is the Manhattan plot for the unadjusted p-values. The highlighted site is cg22798925. C) Plot C is the QQ-plot for the adjusted p-values using Bacon and Cate. D) Plot D is the Manhattan plot for adjusted p-values using Bacon and Cate. The highlighted site is cg22798925.

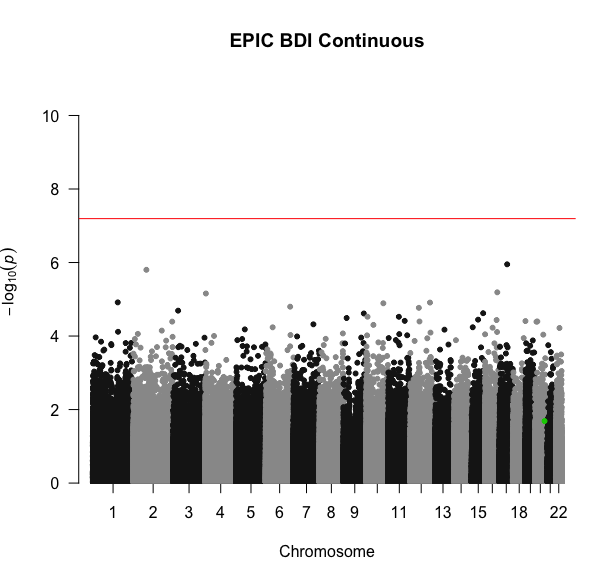

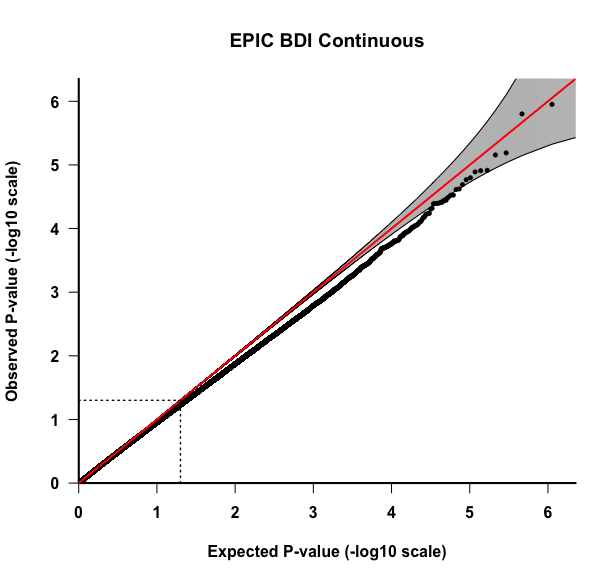

A	B

C	D

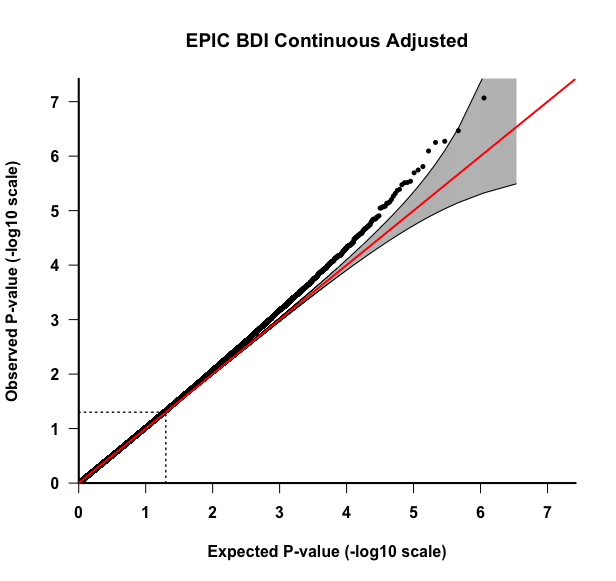

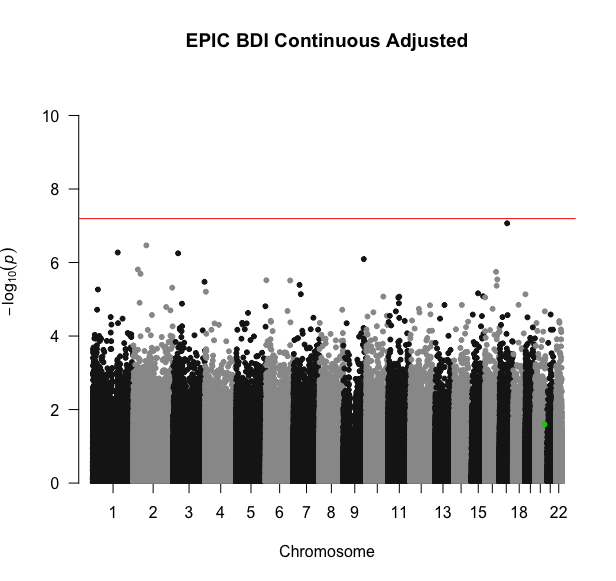

**Figure S4: Results for the EPIC EWAS for the BDI-II continuous variable.** All association models were adjusted for covariates: mother’s smoking status, average household income, child’s sex, preterm birth (<37 weeks), first three cell type PCs, and first five genotype PCs. A) Plot A is the QQ-plot for the unadjusted p-values. B) Plot B is the Manhattan plot for the unadjusted p-values. The highlighted site is cg22798925. C) Plot C is the QQ-plot for the adjusted p-values using Bacon and Cate. D) Plot D is the Manhattan plot for adjusted p-values using Bacon and Cate. The highlighted site is cg22798925.

**S5: Comparison between the p-values and beta estimates for the EPDS and BDI-II**
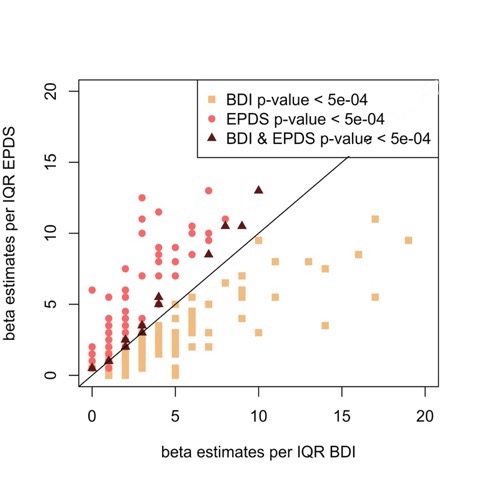

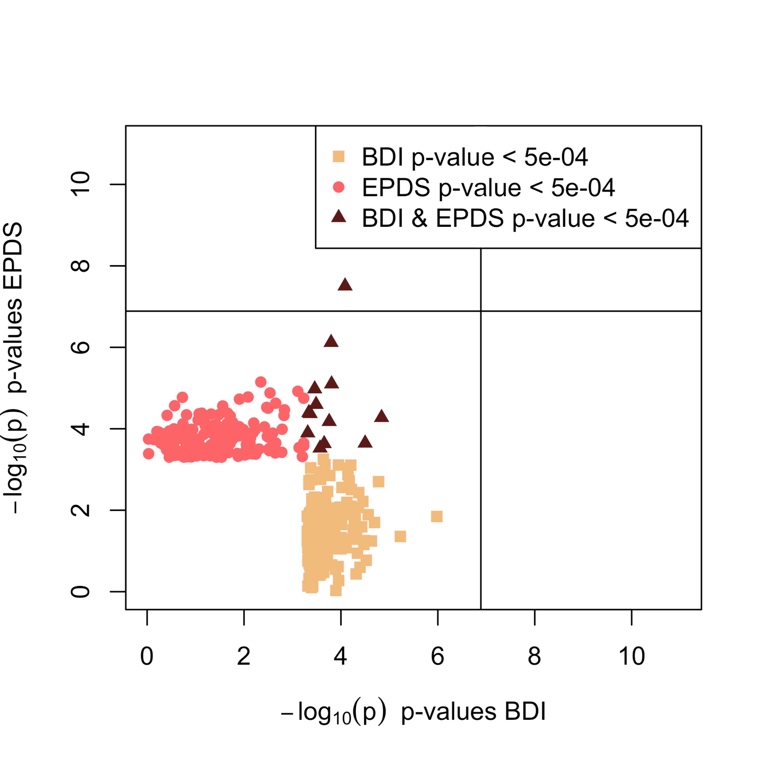
**continuous variables from the meta-analysis.** The meta-analysis was adjusted for mother’s smoking status, average household income, child’s sex, preterm birth (<37 weeks), first three cell type PCs, and first five genotype PCs. Unmeasured confounding and bias were adjusted with Cate and Bacon R packages. A) Plot A is for the p-values between the EPDS and BDI-II continuous variables below a threshold of 5e-04. B) Plot B is for the beta estimates per IQR for the EPDS and BDI-II continuous variables. The plotted values are the beta estimates divided by the IQR to account for the different ranges between the EPDS and BDI-II scales.

B

A

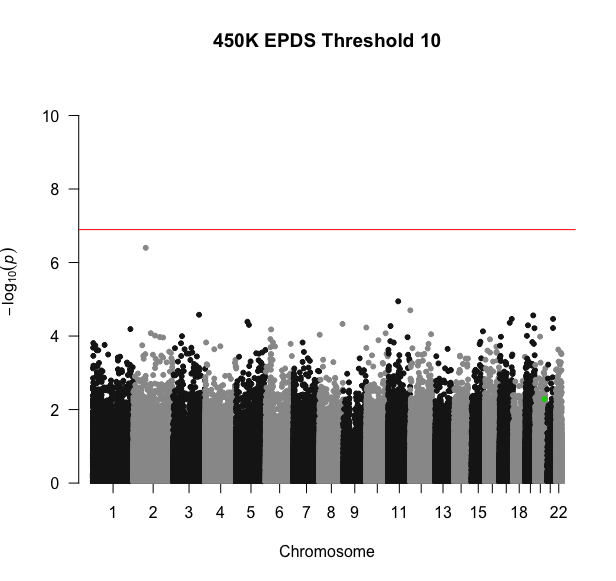

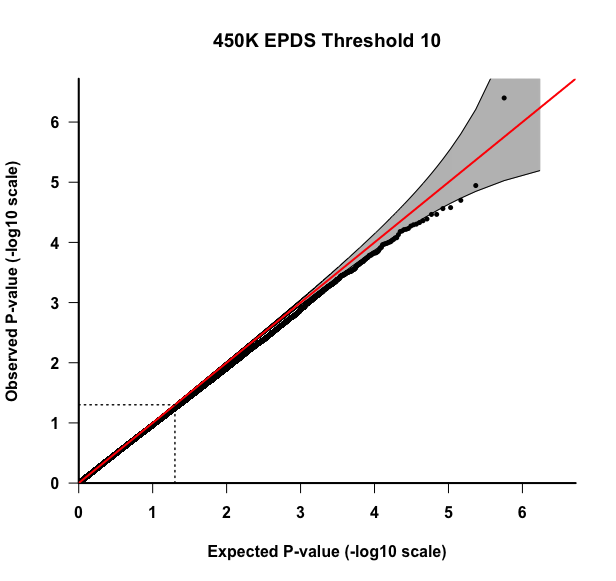

A	B

C	D

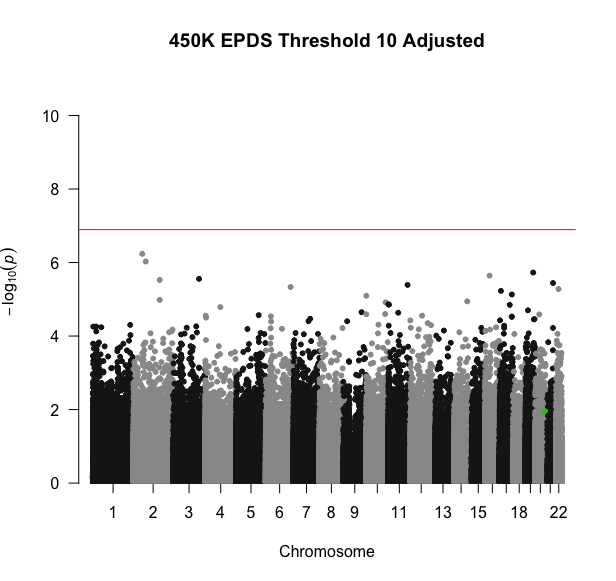

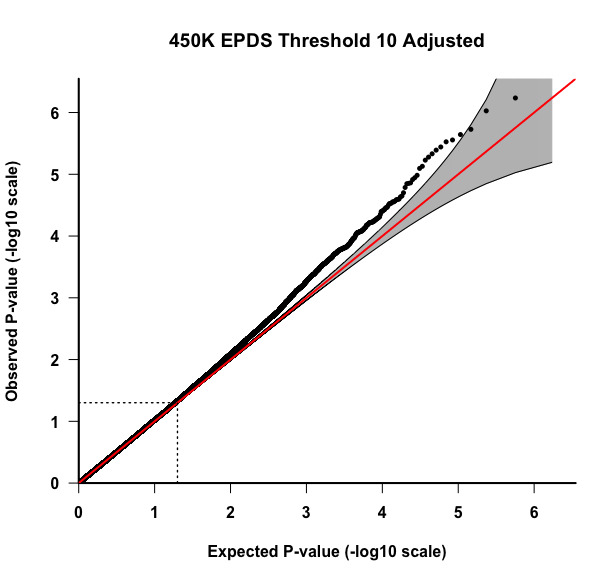

**Figure S6: Results for the 450K EWAS for the EPDS 10 threshold variable.** All association models were adjusted for covariates: mother’s smoking status, average household income, child’s sex, preterm birth (<37 weeks), first three cell type PCs, and first five genotype PCs. A) Plot A is the QQ-plot for the unadjusted p-values. B) Plot B is the Manhattan plot for the unadjusted p-values. The highlighted site is cg22798925. C) Plot C is the QQ-plot for the adjusted p-values using Bacon and Cate. D) Plot D is the Manhattan plot for adjusted p-values using Bacon and Cate. The highlighted site is cg22798925.

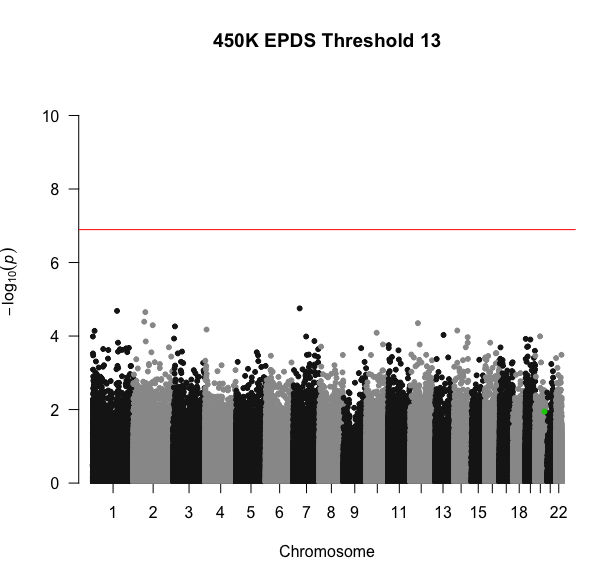

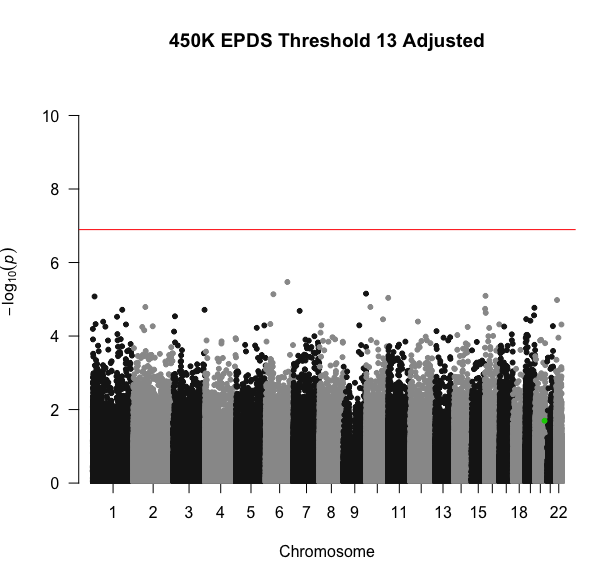

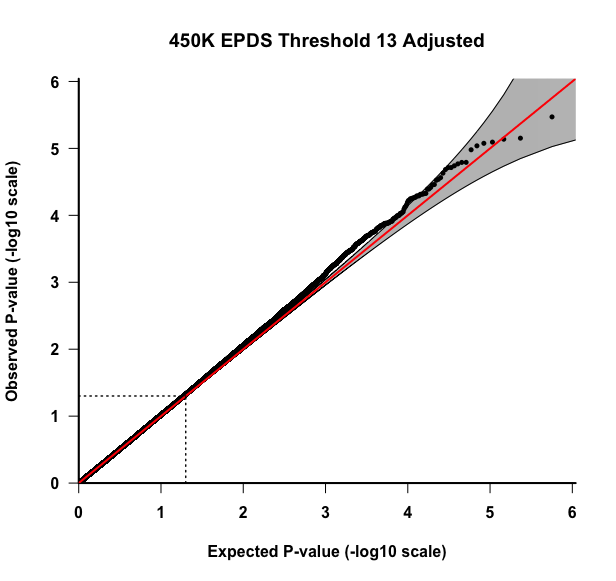

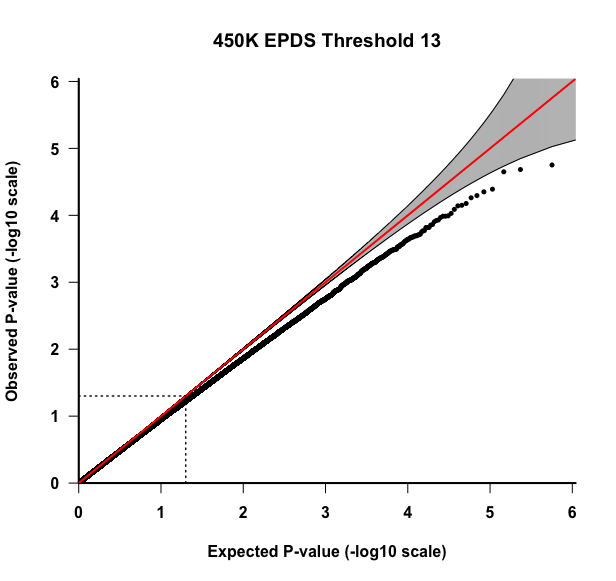
**Figure S7: Results for the 450K EWAS for the BDI-II 20 threshold variable.** All association models were adjusted for covariates: mother’s smoking status, average household income, child’s sex, preterm birth (<37 weeks), first three cell type PCs, and first five genotype PCs. A) Plot A is the QQ-plot for the unadjusted p-values. B) Plot B is the Manhattan plot for the unadjusted p-values. The highlighted site is cg22798925. C) Plot C is the QQ-plot for the adjusted p-values using Bacon and Cate. D) Plot D is the Manhattan plot for adjusted p-values using Bacon and Cate. The highlighted site is cg22798925.

A	B

C	D

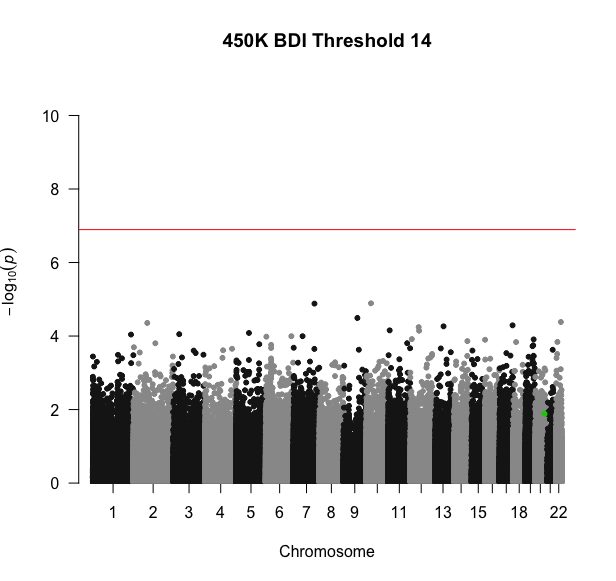

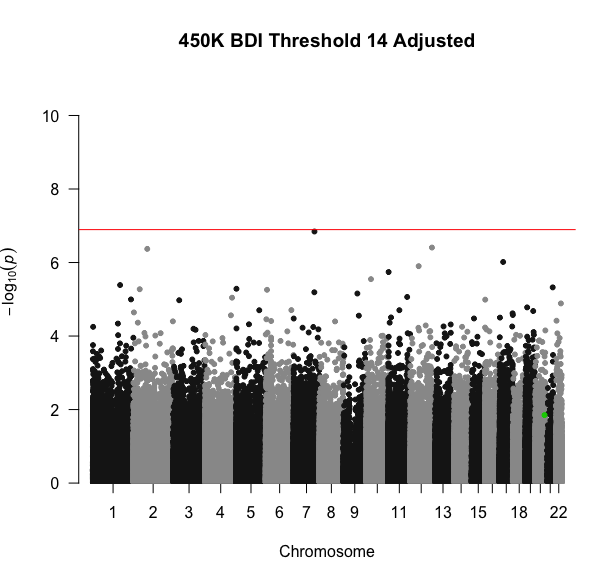

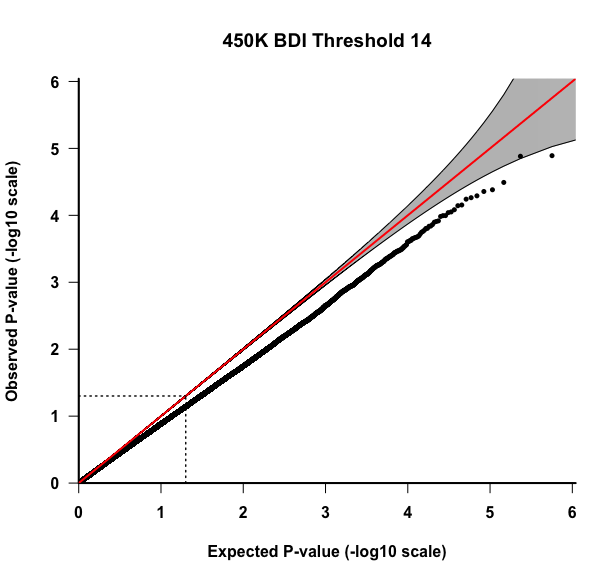

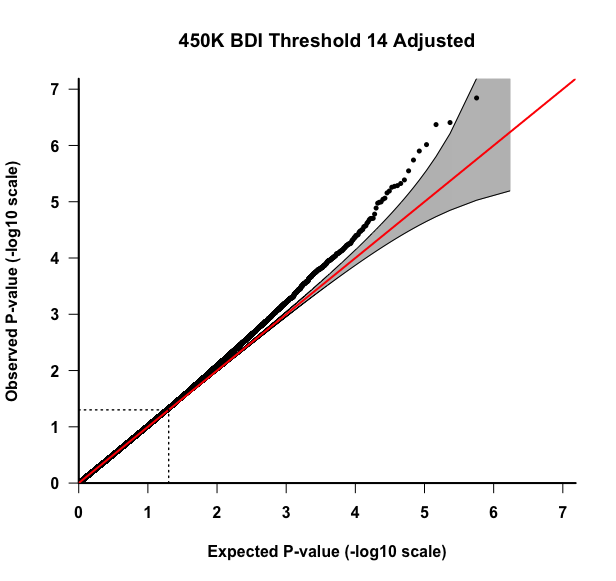

A	B

C	D

**Figure S8: Results for the 450K EWAS for the BDI-II 14 threshold variable.** All association models were adjusted for covariates: mother’s smoking status, average household income, child’s sex, preterm birth (<37 weeks), first three cell type PCs, and first five genotype PCs. A) Plot A is the QQ-plot for the unadjusted p-values. B) Plot B is the Manhattan plot for the unadjusted p-values. The highlighted site is cg22798925. C) Plot C is the QQ-plot for the adjusted p-values using Bacon and Cate. D) Plot D is the Manhattan plot for adjusted p-values using Bacon and Cate. The highlighted site is cg22798925.

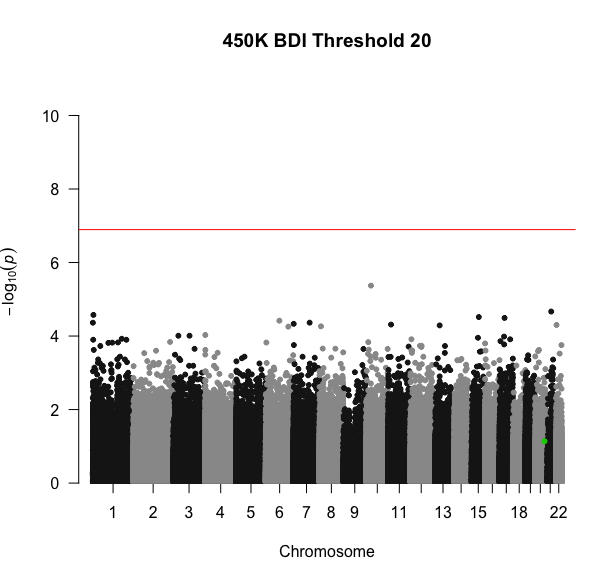

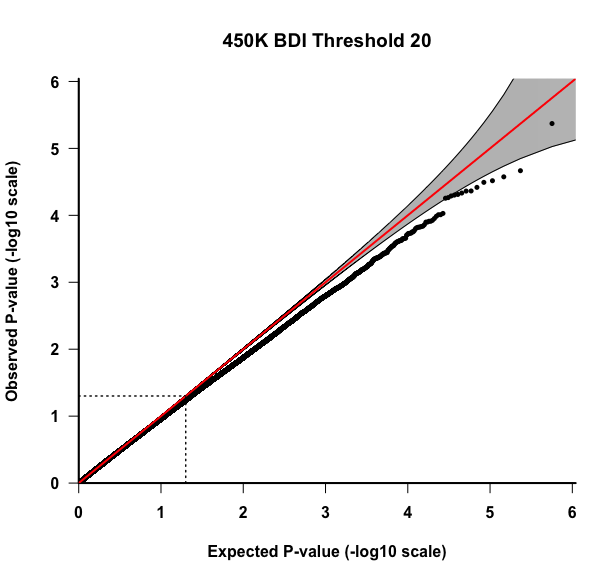

A	B

C	D

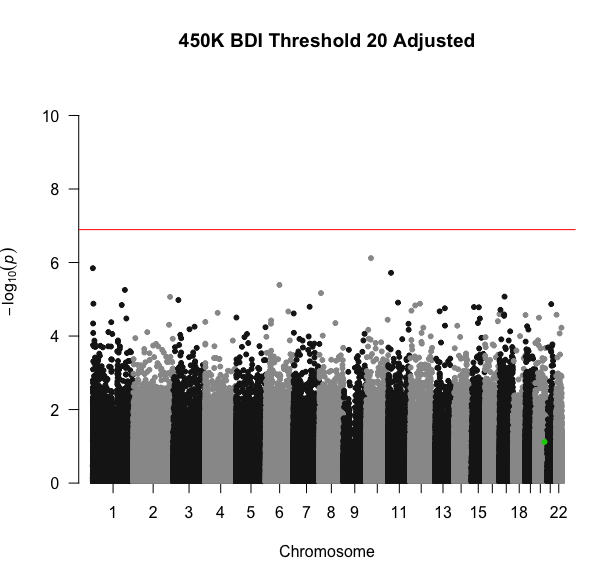

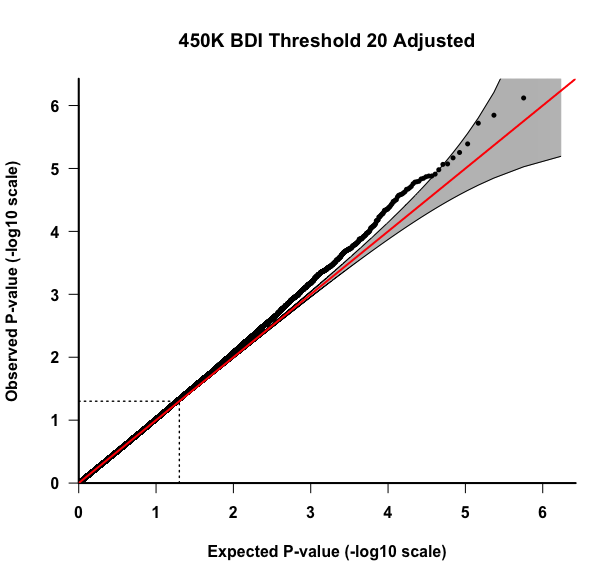

**Figure S9: Results for the 450K EWAS for the BDI-II 20 threshold variable.** All association models were adjusted for covariates: mother’s smoking status, average household income, child’s sex, preterm birth (<37 weeks), first three cell type PCs, and first five genotype PCs. A) Plot A is the QQ-plot for the unadjusted p-values. B) Plot B is the Manhattan plot for the unadjusted p-values. The highlighted site is cg22798925. C) Plot C is the QQ-plot for the adjusted p-values using Bacon and Cate. D) Plot D is the Manhattan plot for adjusted p-values using Bacon and Cate. The highlighted site is cg22798925.

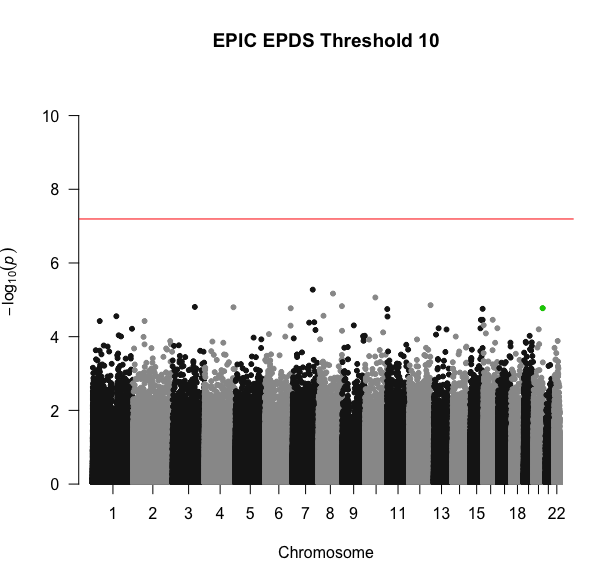

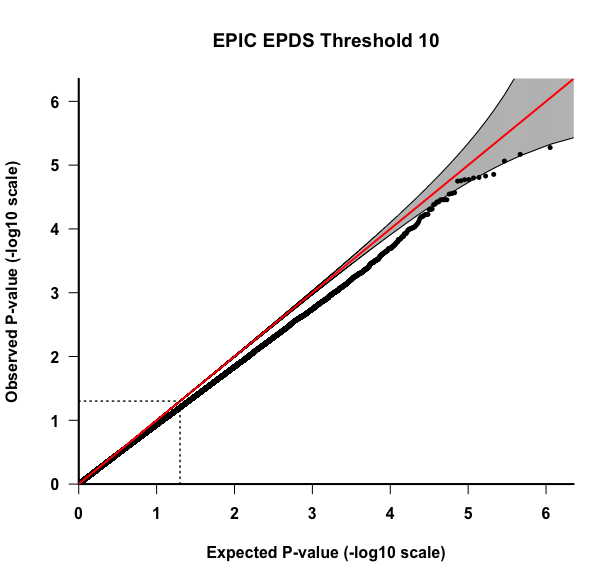

A	B

C	D

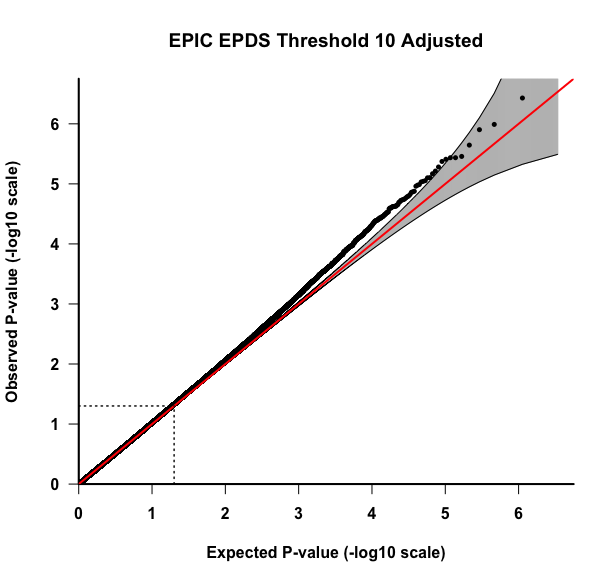

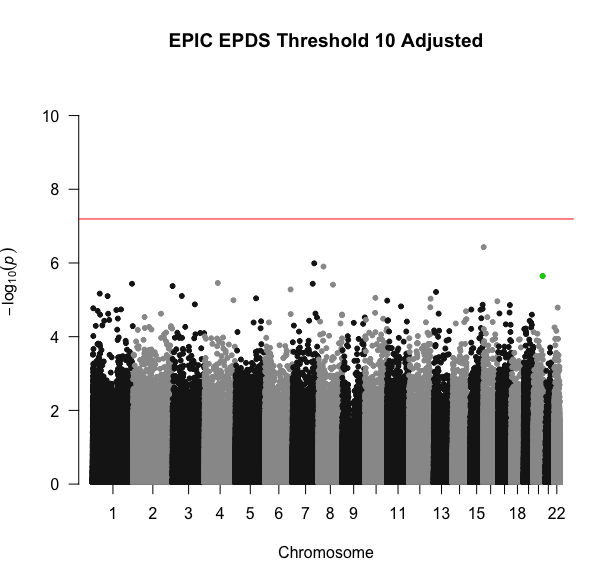

**Figure S10: Results for the EPIC EWAS for the EPDS 10 threshold variable.** All association models were adjusted for covariates: mother’s smoking status, average household income, child’s sex, preterm birth (<37 weeks), first three cell type PCs, and first five genotype PCs. A) Plot A is the QQ-plot for the unadjusted p-values. B) Plot B is the Manhattan plot for the unadjusted p-values. The highlighted site is cg22798925. C) Plot C is the QQ-plot for the adjusted p-values using Bacon and Cate. D) Plot D is the Manhattan plot for adjusted p-values using Bacon and Cate. The highlighted site is cg22798925.

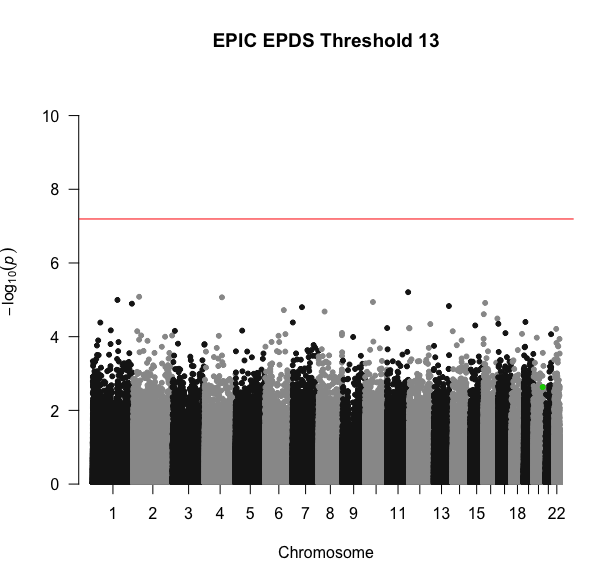

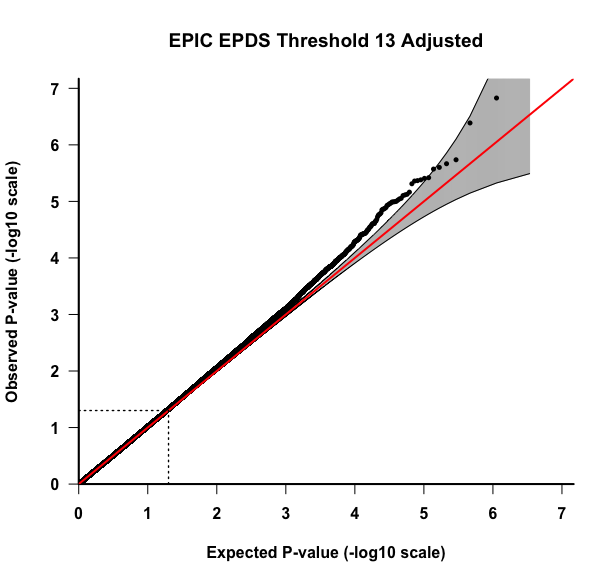

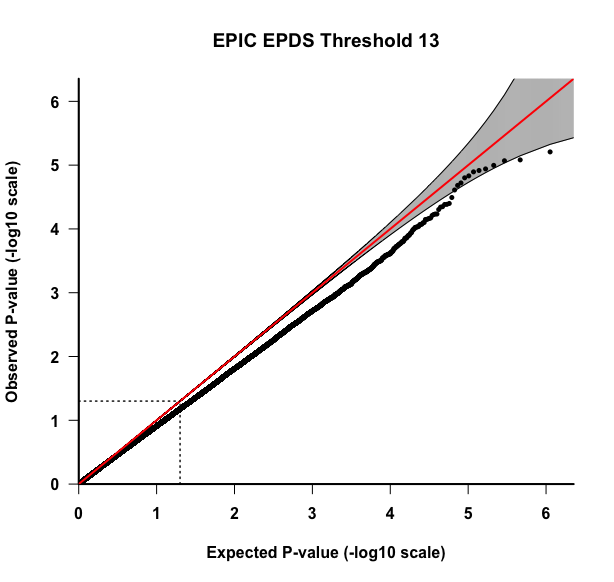

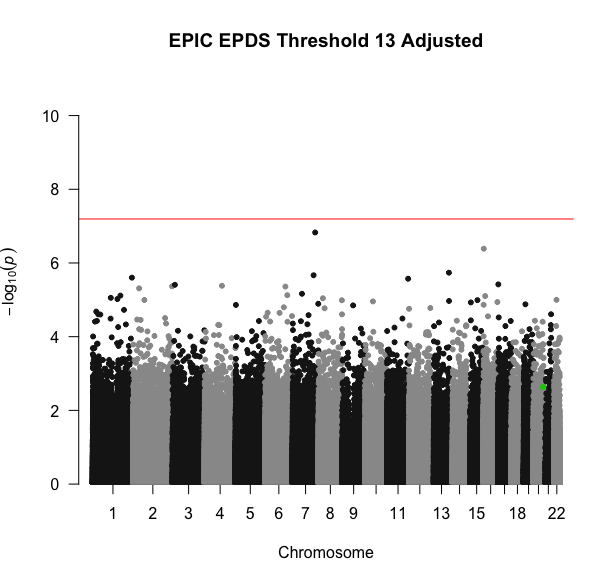

A	B

C	D

**Figure S11: Results for the EPIC EWAS for the EPDS 13 threshold variable.** All association models were adjusted for covariates: mother’s smoking status, average household income, child’s sex, preterm birth (<37 weeks), first three cell type PCs, and first five genotype PCs. A) Plot A is the QQ-plot for the unadjusted p-values. B) Plot B is the Manhattan plot for the unadjusted p-values. The highlighted site is cg22798925. C) Plot C is the QQ-plot for the adjusted p-values using Bacon and Cate. D) Plot D is the Manhattan plot for adjusted p-values using Bacon and Cate. The highlighted site is cg22798925.

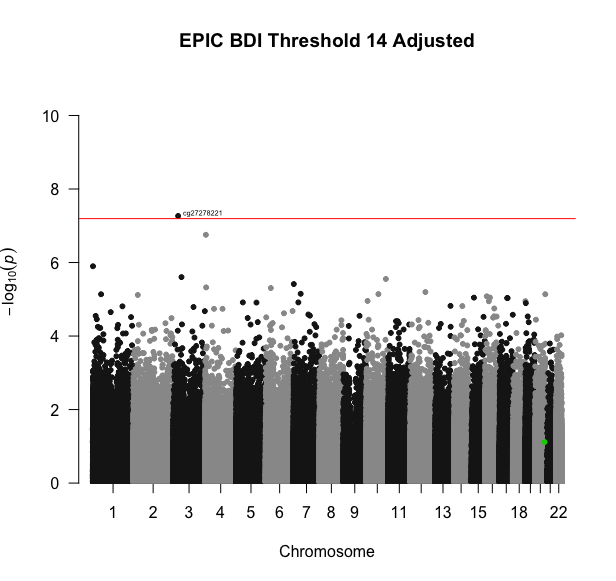

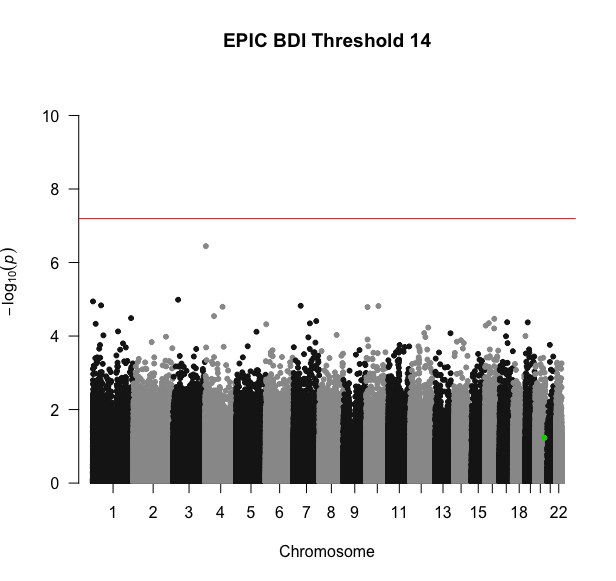

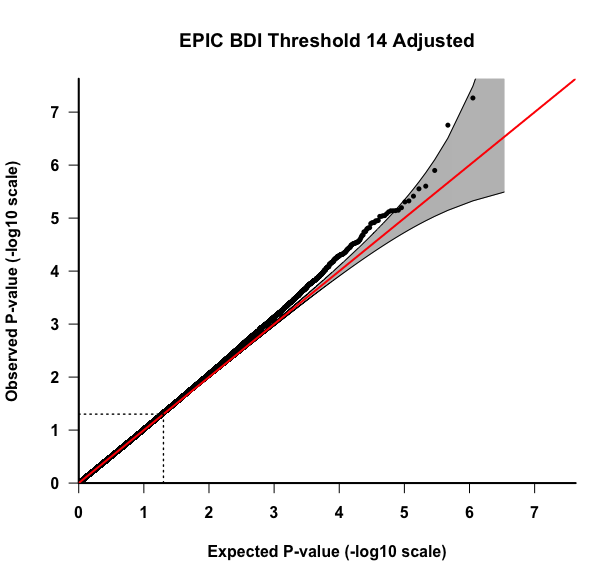

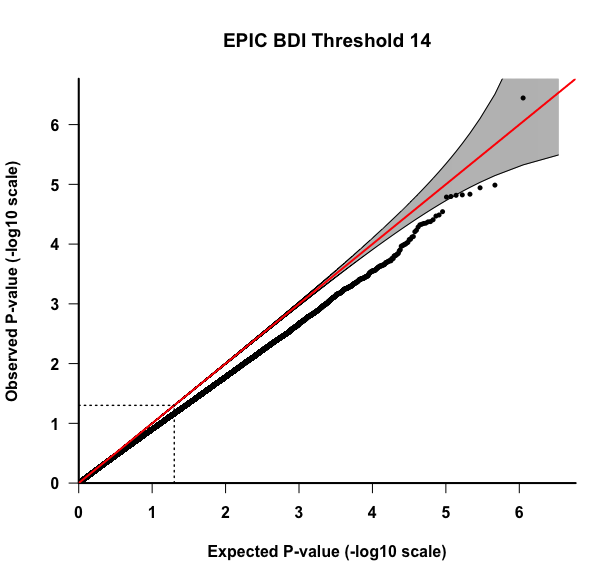
**Figure S12: Results for the EPIC EWAS for the BDI-II 14 threshold variable.** All association models were adjusted for covariates: mother’s smoking status, average household income, child’s sex, preterm birth (<37 weeks), first three cell type PCs, and first five genotype PCs. A) Plot A is the QQ-plot for the unadjusted p-values. B) Plot B is the Manhattan plot for the unadjusted p-values. The highlighted site is cg22798925. C) Plot C is the QQ-plot for the adjusted p-values using Bacon and Cate. D) Plot D is the Manhattan plot for adjusted p-values using Bacon and Cate. The highlighted site is cg22798925.

A	B

C	D

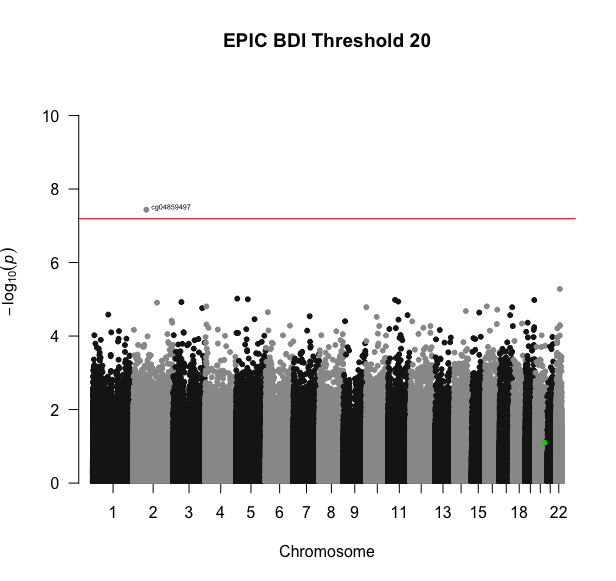

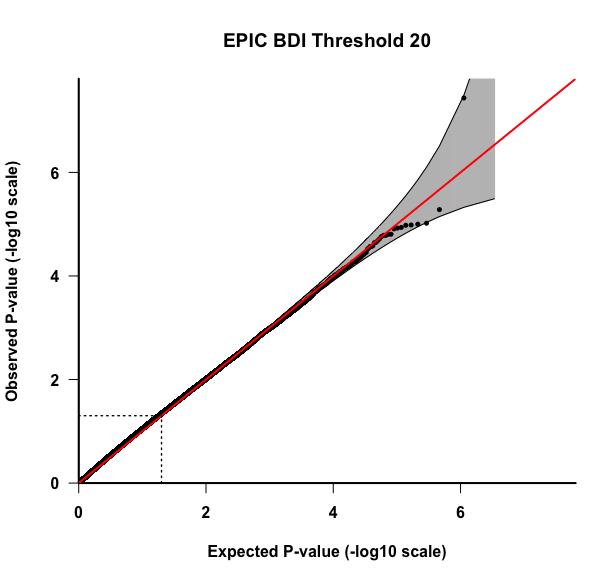

A	B

C	D

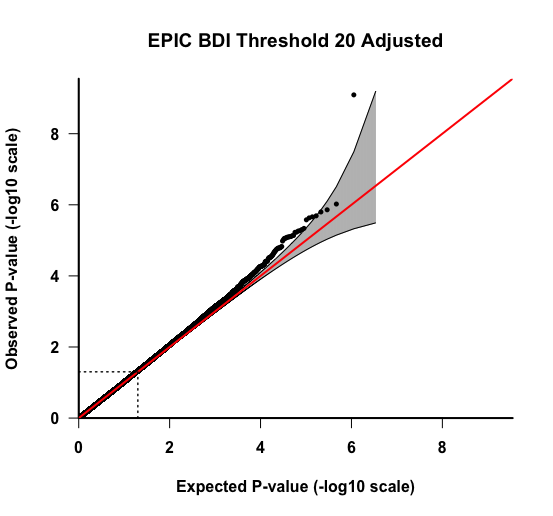

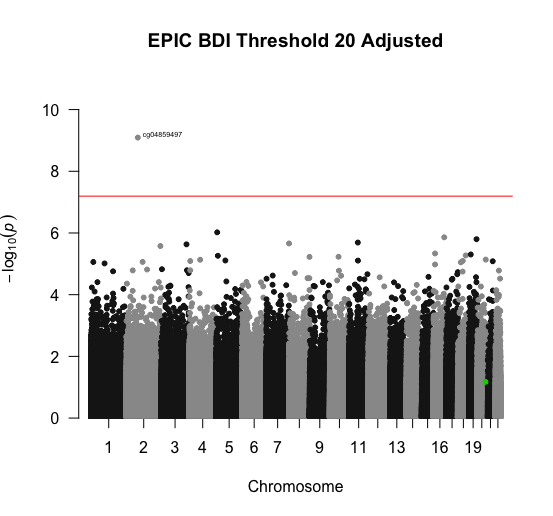

**Figure S13: Results for the EPIC EWAS for the BDI-II 20 threshold variable.** All association models were adjusted for covariates: mother’s smoking status, average household income, child’s sex, preterm birth (<37 weeks), first three cell type PCs, and first five genotype PCs. A) Plot A is the QQ-plot for the unadjusted p-values. B) Plot B is the Manhattan plot for the unadjusted p-values. The highlighted site is cg22798925. C) Plot C is the QQ-plot for the adjusted p-values using Bacon and Cate. D) Plot D is the Manhattan plot for adjusted p-values using Bacon and Cate. The highlighted site is cg22798925.

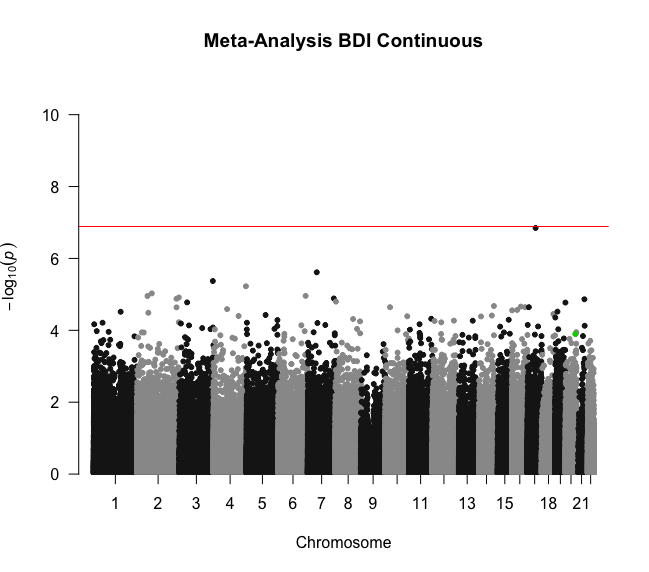

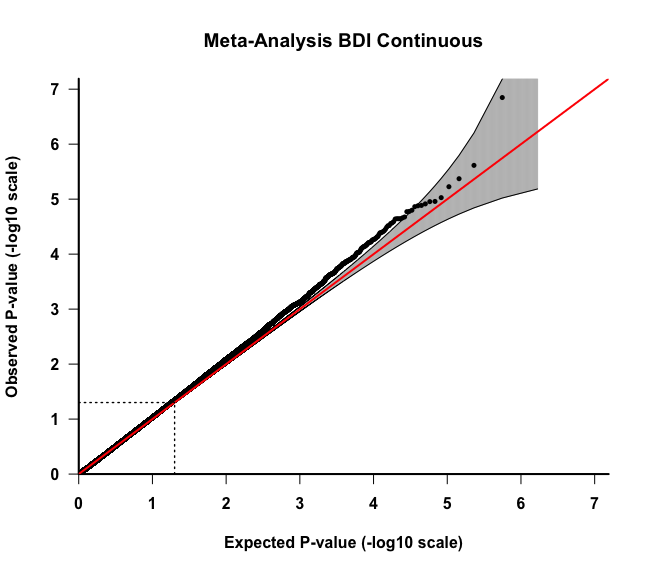
**Figure S14: Results for the meta-analysis for the BDI-II continuous variable.** The meta-analysis was adjusted for all covariates (mother’s smoking status, average household income, child’s sex, preterm birth (<37 weeks), first three cell type PCs, and first five genotype PCs) and the p-values were adjusted for unmeasured confounding using Bacon and Cate prior to the meta-analysis. A) Plot A is the QQ-plot for the p-values. B) Plot B is the Manhattan plot for the p-values. The highlighted site is cg22798925.

A

B

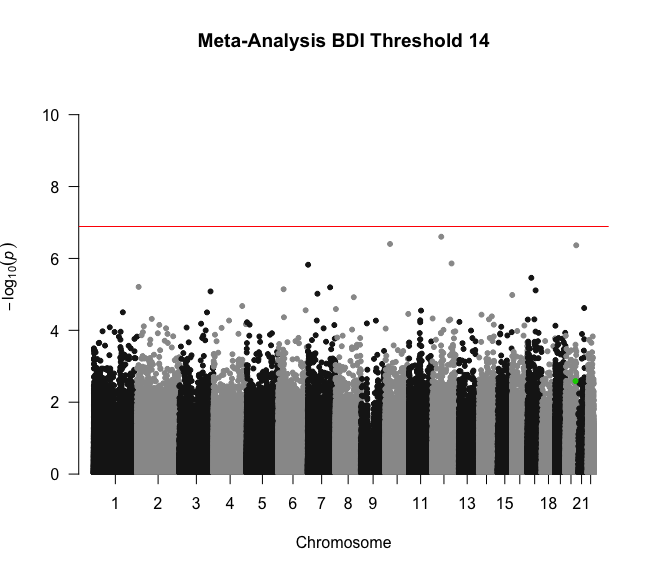

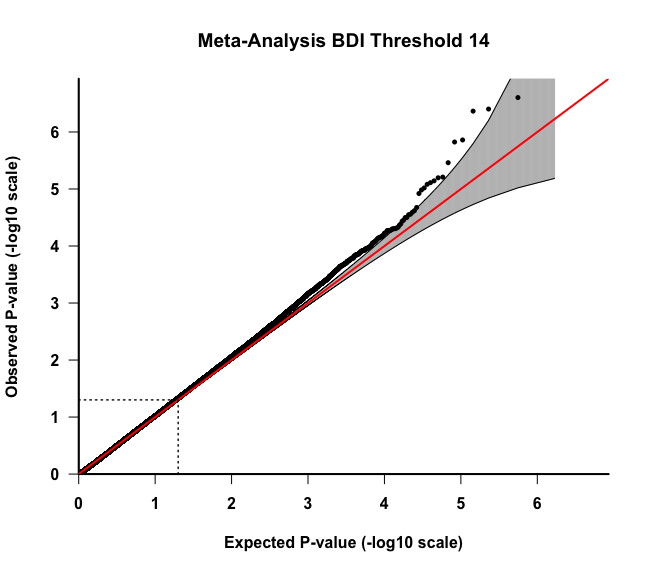

B

A

**Figure S15: Results for the meta-analysis for the BDI-II 14 threshold variable.** The meta-analysis was adjusted for all covariates (mother’s smoking status, average household income, child’s sex, preterm birth (<37 weeks), first three cell type PCs, and first five genotype PCs) and the p-values were adjusted for unmeasured confounding using Bacon and Cate prior to the meta-analysis. A) Plot A is the QQ-plot for the p-values. B) Plot B is the Manhattan plot for the p-values. The highlighted site is cg22798925.

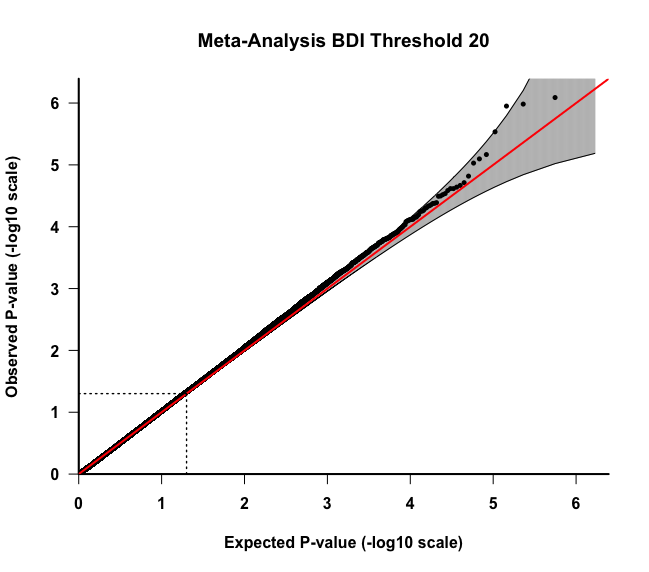

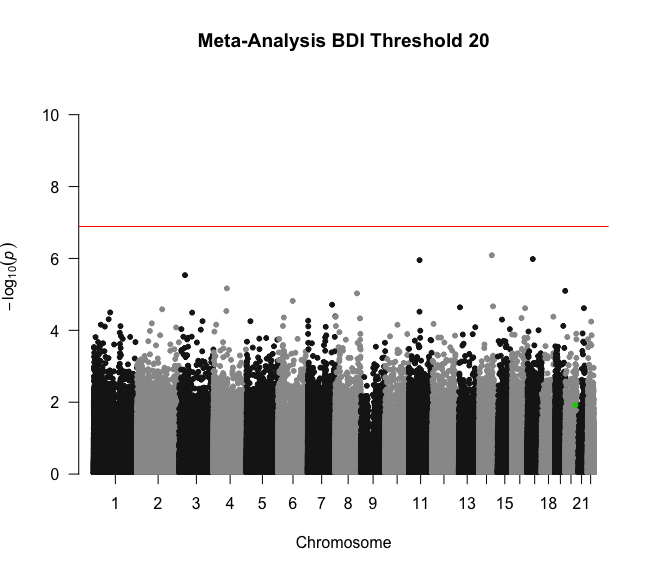

A

B

**Figure S16: Results for the meta-analysis for the BDI-II 20 threshold variable.** The meta-analysis was adjusted for all covariates (mother’s smoking status, average household income, child’s sex, preterm birth (<37 weeks), first three cell type PCs, and first five genotype PCs) and the p-values were adjusted for unmeasured confounding using Bacon and Cate prior to the meta-analysis. A) Plot A is the QQ-plot for the p-values. B) Plot B is the Manhattan plot for the p-values. The highlighted site is cg22798925.

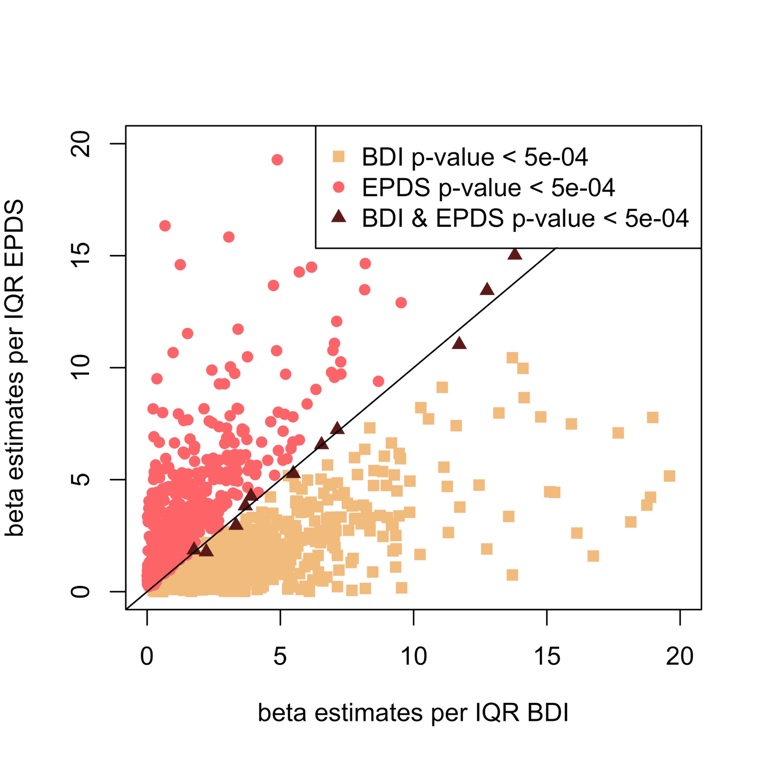

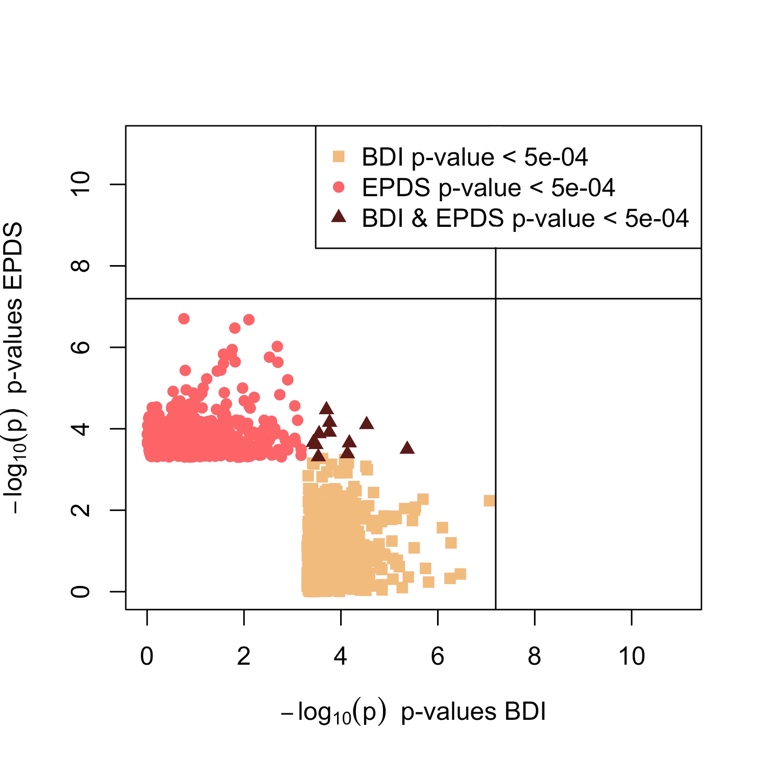
**Figure S17: Comparison between the p-values and beta estimates for the EPDS and BDI-II continuous variables from the EPIC EWAS.** All association models were adjusted for covariates: mother’s smoking status, average household income, child’s sex, preterm birth (<37 weeks), first three cell type PCs, and first five genotype PCs. Unmeasured confounding and bias were adjusted with Cate and Bacon R packages. A) Plot A is for the p-values between the EPDS and BDI-II continuous variables below a threshold of 5e-04. B) Plot B is for the beta estimates per IQR for the EPDS and BDI-II continuous variables. The plotted values are the beta estimates divided by the IQR to account for the different ranges between the EPDS and BDI-II scales.

A

B

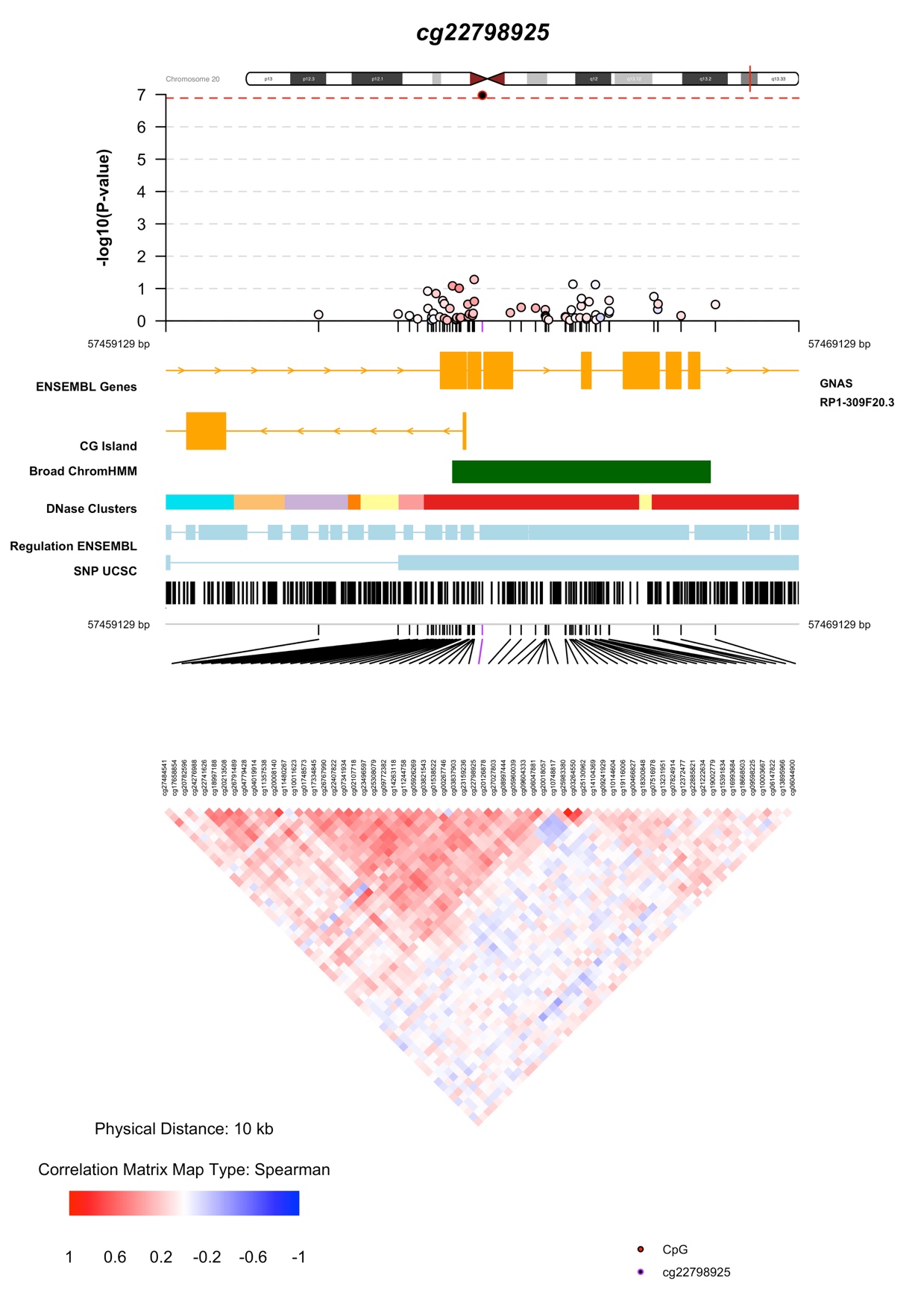

**Figure S18: CoMET results for cg22798925.** The CoMET results were obtained using p-values from the meta-analysis for the EPIC continuous variable while adjusting for all covariates. The CpG sites include sites 5000 bp upstream and downstream cg22798925.

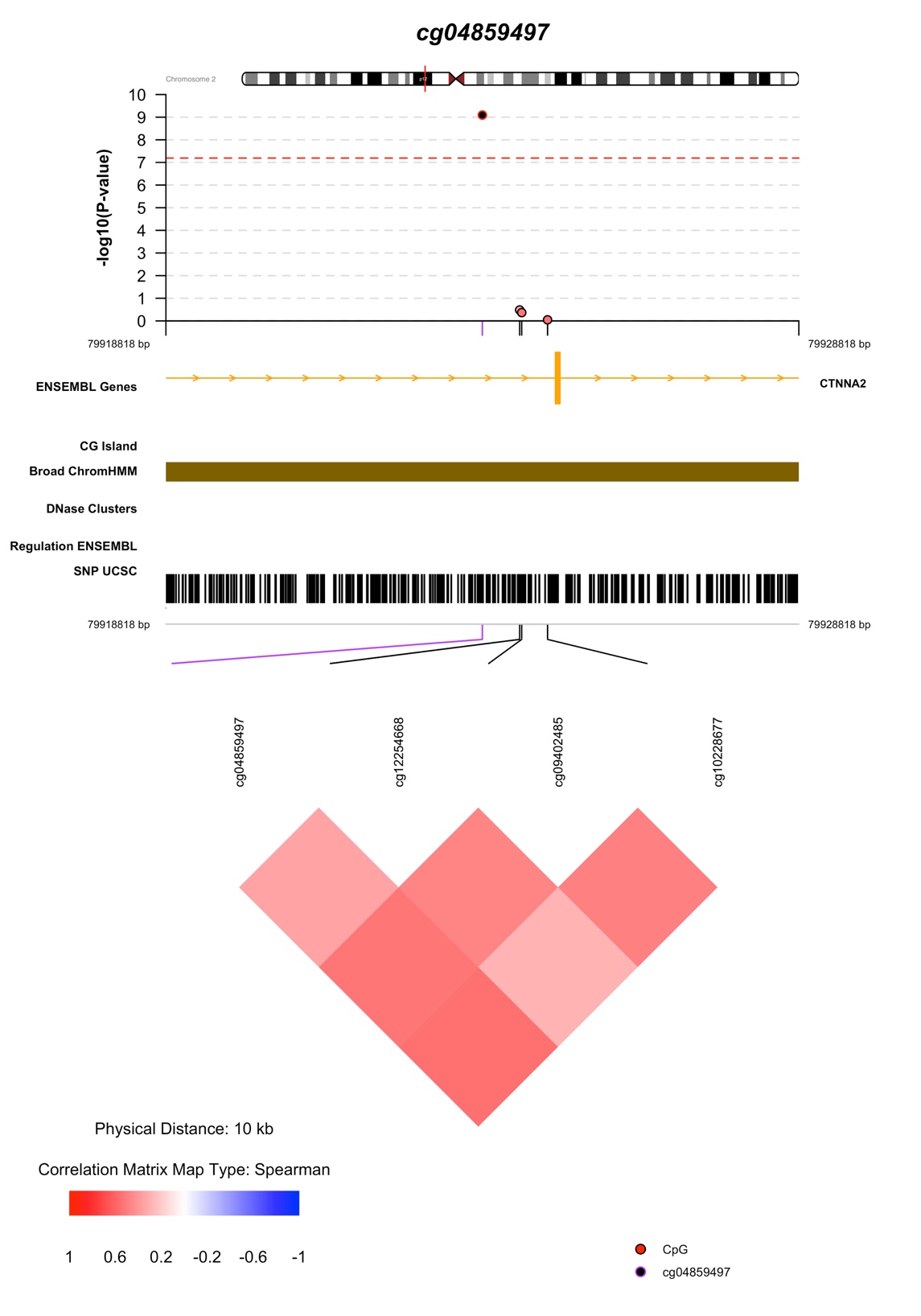

**Figure S19: CoMET results for cg04859497.** The CoMET results were obtained using p-values from the EPIC EWAS for the BDI-II 20 threshold variable while adjusting for all covariates. The CpG sites include sites 5000 bp upstream and downstream cg04859497.

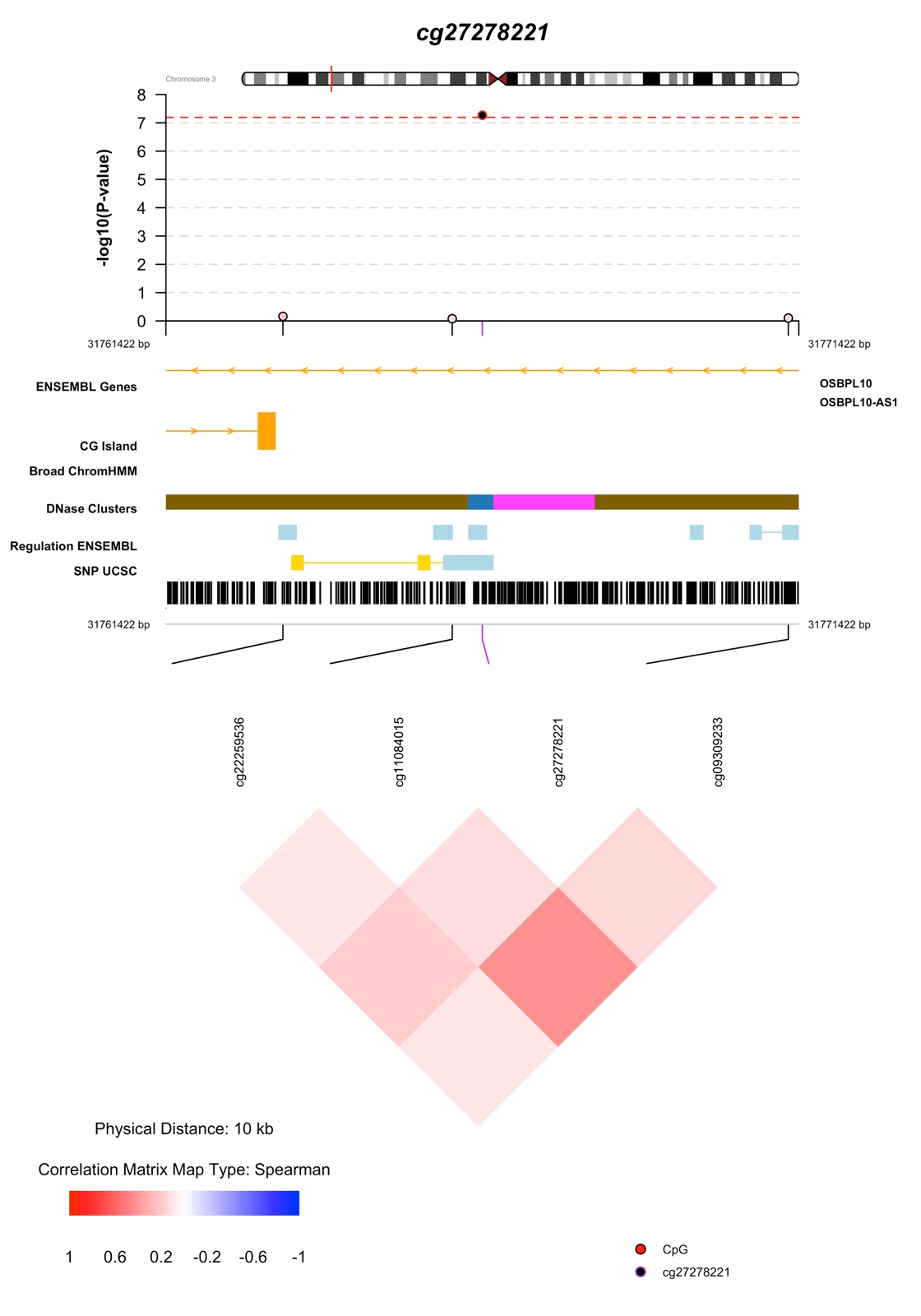

**Figure S20: CoMET results for cg27278221.** The CoMET results were obtained using p-values from the EPIC EWAS for the BDI-II 14 threshold variable while adjusting for all covariates. The CpG sites include sites 5000 bp upstream and downstream cg27278221.

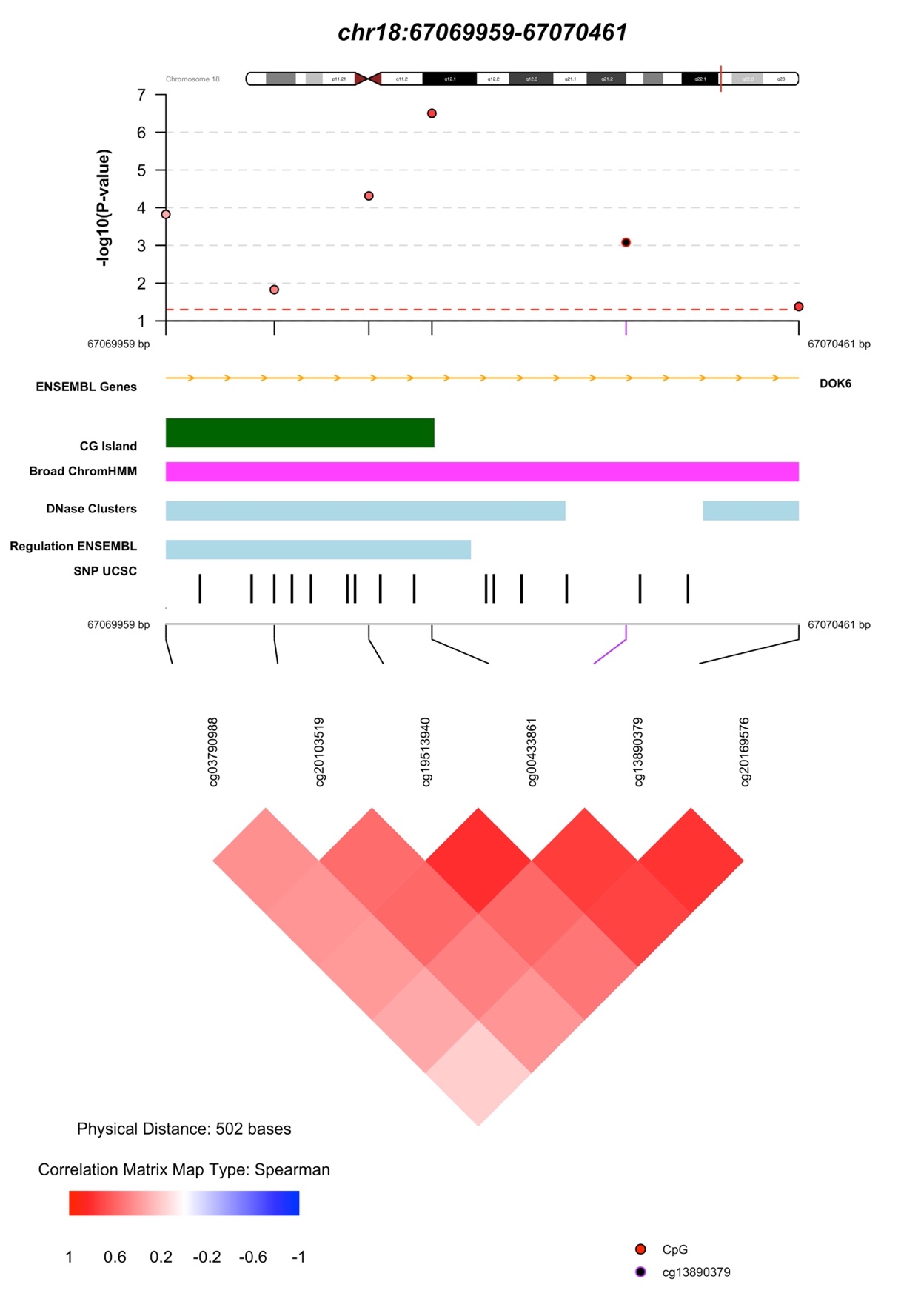

**Figure S21: CoMET results for DMR chr18:67069959-67070461.** The CoMET results were obtained using p-values from the meta-analysis for the EPDS threshold-13 variable while adjusting for all covariates.

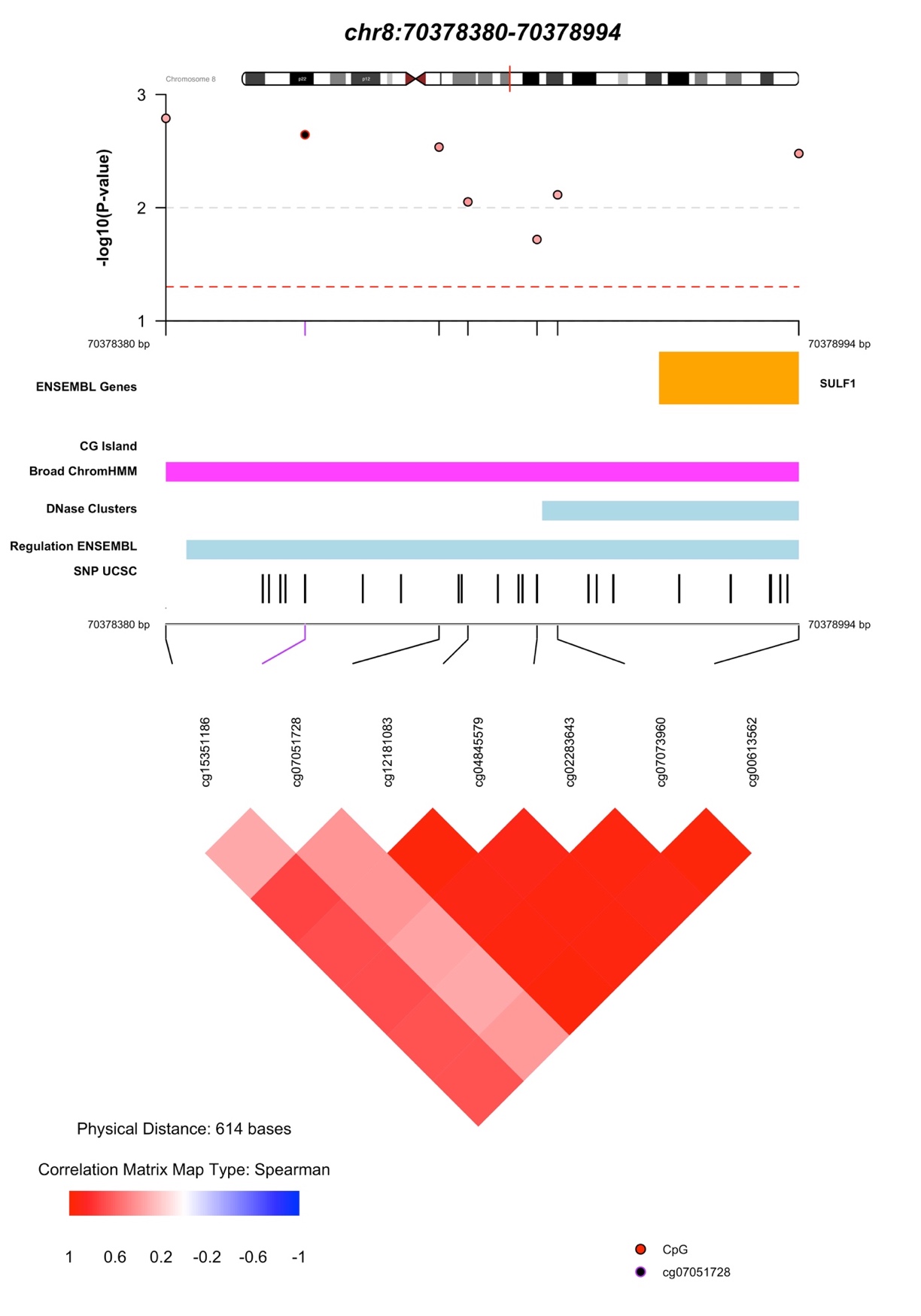

**Figure S22: CoMET results for DMR chr8:70378380-70378994.** The CoMET results were obtained using p-values from the meta-analysis for the BDI-II threshold-20 variable while adjusting for all covariates.

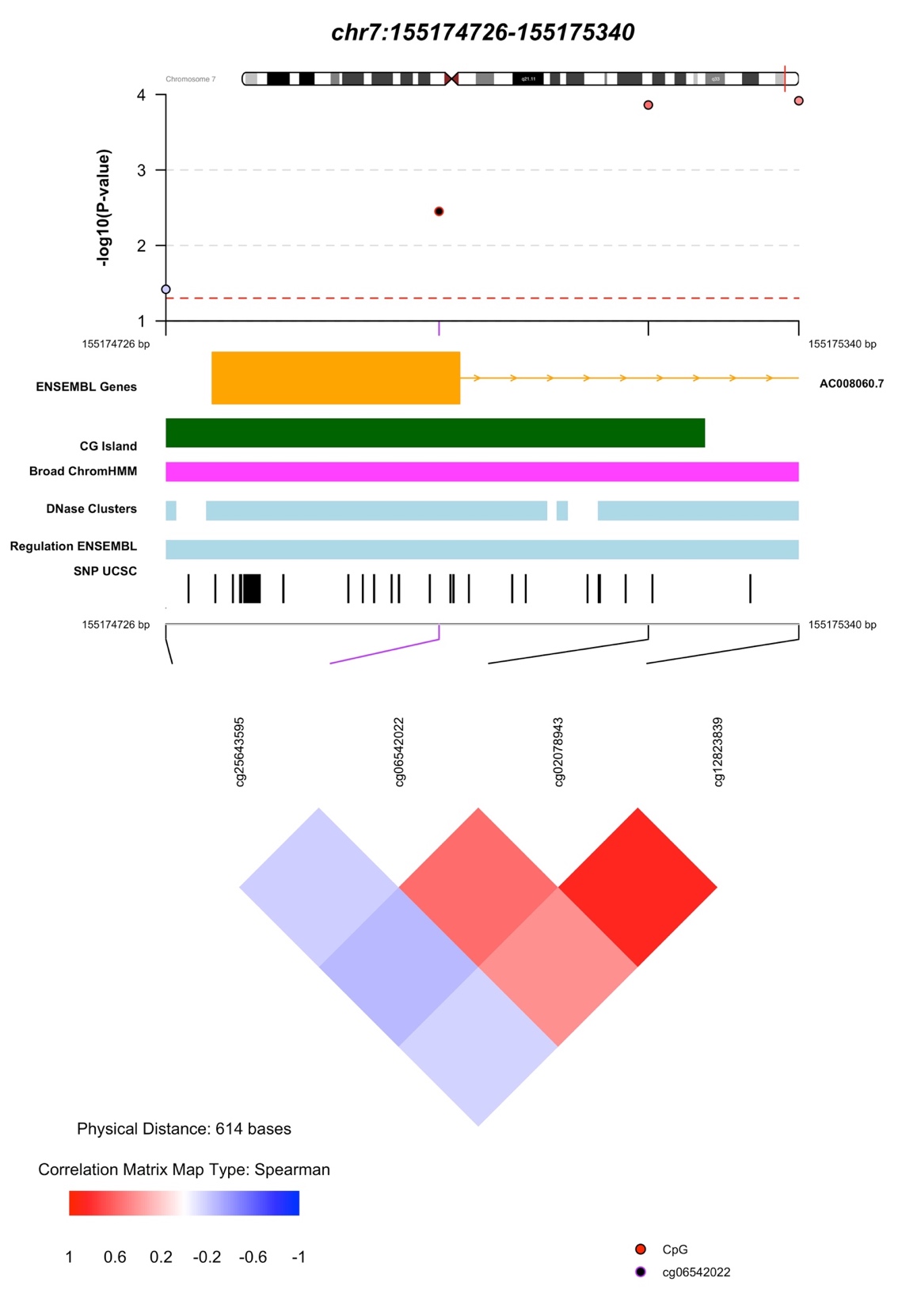

**Figure S23: CoMET results for DMR chr7:155174726-155175340.** The CoMET results were obtained using p-values from the meta-analysis for the BDI-II continuous variable while adjusting for all covariates.

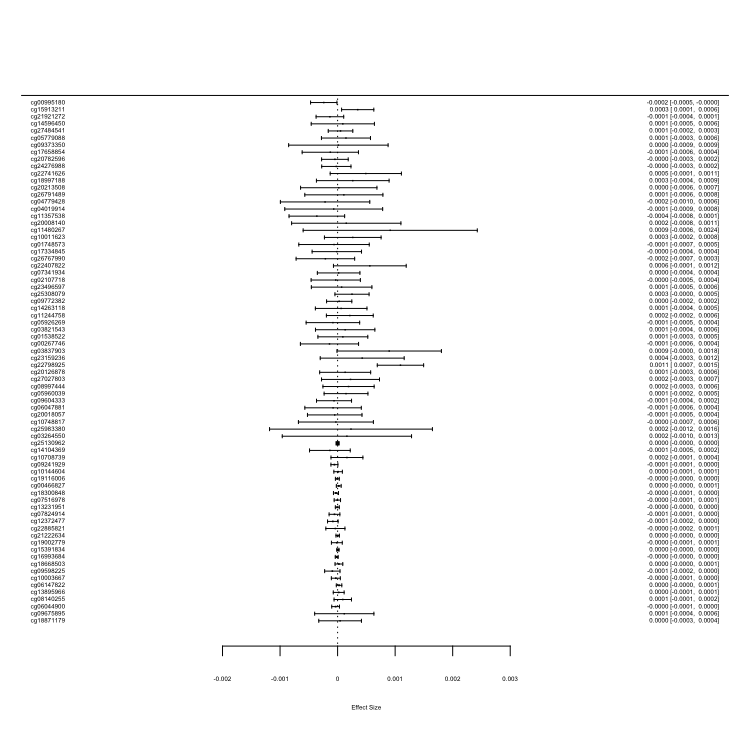

Effect Size per IQR

[95% CI]

CpG Site

**Figure S24: Effect size per IQR for CpG sites surrounding cg22798925.** CpG sites include sites 5000 bps upstream and downstream cg22798925. Effect sizes were obtained through the meta-analysis for the EPDS continuous depression variable.

**Supplementary Tables**

**Table S1: DMR results for meta-analysis EPDS continuous variable**

| DMR | # CpGs | P-value | Max Effect | Overlapping Promoters |
| --- | --- | --- | --- | --- |
| chr1: 201951162-201951234 | 2 | 4.30E-03 | 0.0013 | RNPEP-001, RNPEP-003, RNPEP-008, RNPEP-002, RNPEP-007, RNPEP-010, RNPEP-004, RNPEP-005, RNPEP-006, RNPEP-009 |
| chr1: 55035183-155035748 | 5 | 1.37E-03 | -0.0012 | EFNA4-002, EFNA4-001, EFNA4-003, ADAM15-035, EFNA3-001, EFNA3-202 |
| chr10: 45406187-45406847 | 5 | 2.08E-04 | 0.0062 | TMEM72-001, TMEM72-201 |
| chr10: 91296252-91296457 | 3 | 4.86E-03 | -0.0021 | SLC16A12-201, SLC16A12-001 |
| chr10: 94180383-94180753 | 4 | 4.58E-03 | -0.0013 | MARK2P9-001 |
| chr10: 94820376-94821085 | 5 | 5.96E-03 | -0.0022 | CYP26C1-001, RP11-348J12.2-001 |
| chr11: 111249659-111250201 | 5 | 7.95E-03 | -0.0022 | POU2AF1-001, POU2AF1-002, POU2AF1-003 |
| chr13: 47472050-47472429 | 11 | 5.96E-03 | -0.0018 | HTR2A-001, HTR2A-201, HTR2A-202 |
| chr14: 24422368-24423864 | 12 | 4.43E-08 | -0.0019 | DHRS4-201, DHRS4-002, DHRS4-202, DHRS4-203, DHRS4-001, DHRS4-003, DHRS4-AS1-004, DHRS4-204, DHRS4-004, DHRS4-AS1-003, DHRS4-AS1-005, DHRS4-AS1-002, DHRS4-AS1-007, DHRS4-005, DHRS4-007, DHRS4-006 |
| chr15: 98195808-98196247 | 4 | 1.11E-05 | -0.0032 | NA |
| chr17: 46019006-46019184 | 5 | 7.14E-03 | 0.0005 | PNPO-001, AC003665.1-003, AC003665.1-002, PNPO-002, AC003665.1-001, PNPO-201, PNPO-202, PNPO-003, PNPO-004, PNPO-008, PNPO-005, AC003665.1-004, PNPO-007 |
| chr17: 6899085-6899888 | 12 | 1.36E-04 | -0.0033 | ALOX12-001, ALOX12-003, RP11-589P10.5-001 |
| chr18: 67069959-67070461 | 6 | 4.41E-10 | -0.0026 | DOK6-001 |
| chr19: 18698825-18699631 | 9 | 1.46E-05 | -0.0034 | C19orf60-001, C19orf60-002, C19orf60-007, C19orf60-005, C19orf60-008, C19orf60-004, C19orf60-003, C19orf60-006 |
| chr2:1 08994116-108994528 | 5 | 2.46E-03 | 0.0019 | SULT1C4-001, SULT1C4-004, SULT1C4-003 |
| chr4: 74847710-74848016 | 7 | 7.14E-03 | 0.0028 | PF4-001 |
| chr6: 33282313-33283317 | 27 | 1.80E-08 | -0.0016 | TAPBP-008, TAPBP-001, TAPBP-003, TAPBP-209, TAPBP-010, TAPBP-007, TAPBP-004, TAPBP-002, TAPBP-006 |
| chr7: 116139180-116139705 | 9 | 2.73E-04 | 0.0018 | CAV2-001, CAV2-019, CAV2-023, AC002066.1-003, CAV2-002 |
| chr8: 142183179-142183860 | 5 | 7.25E-04 | 0.0021 | DENND3-019 |
| chr8: 143085750-143085905 | 2 | 6.29E-03 | 0.0014 | NA |
| chr8: 22132874-22133356 | 7 | 5.96E-03 | 0.0021 | PIWIL2-001, PIWIL2-003, PIWIL2-002, CTD-2530N21.4-001 |

**Table S2: DMR results for meta-analysis BDI-II continuous variable**

| DMRs | # CpGs | P-value | Max Effect | Overlapping Promoters |
| --- | --- | --- | --- | --- |
| chr10: 70321574-70322442 | 7 | 7.86E-05 | 1.58E-03 | TET1-001 |
| chr10: 94820376-94820923 | 3 | 4.03E-03 | -9.66E-04 | CYP26C1-001, RP11-348J12.2-001 |
| chr11: 65190825-65191707 | 4 | 2.74E-04 | 2.50E-03 | NEAT1-002, NEAT1-001, NEAT1-202 |
| chr12: 7781004-7781431 | 5 | 2.16E-03 | -3.85E-03 | NA |
| chr12: 8380050-8380472 | 5 | 4.71E-03 | 1.40E-03 | FAM90A1-001, ALG1L10P-001, FAM90A1-003, FAM90A1-002, AC092111.1-201 |
| chr12:104697193-104697983 | 12 | 2.70E-04 | 1.05E-03 | EID3-001 |
| chr13: 113540189-113540557 | 5 | 5.36E-03 | 1.77E-03 | AL356740.1-201 |
| chr18: 67069959-67070461 | 6 | 3.87E-06 | -1.09E-03 | DOK6-001 |
| chr2: 208631259-208631916 | 3 | 1.01E-03 | 9.52E-04 | NA |
| chr2: 240884831-240884925 | 2 | 2.62E-04 | 2.03E-03 | NA |
| chr2: 27530884-27531535 | 8 | 1.43E-03 | -8.26E-04 | UCN-001 |
| chr3: 194119861-194120150 | 4 | 8.60E-03 | 8.51E-04 | GP5-201, GP5-001 |
| chr5: 176046902-176047485 | 3 | 8.09E-04 | 9.21E-04 | NA |
| chr6: 33047944-33049360 | 16 | 1.79E-11 | 2.06E-03 | HLA-DPB1-002, HLA-DPA1-004, HLA-DPA1-001, HLA-DPB1-008, RPL32P1-001, HLA-DPA1-002, HLA-DPB1-006, HLA-DPA1-005, HLA-DPB1-009, HLA-DPB1-005, HLA-DPB1-007 |
| chr7: 155174726-155175340 | 4 | 3.47E-05 | 1.62E-03 | AC008060.7-001 |
| chr8: 70378380-70378994 | 7 | 2.36E-04 | 1.25E-03 | SULF1-201, SULF1-001, SULF1-008, SULF1-009, SULF1-010 |

**Table S3: DMR results for meta-analysis EPDS threshold-10 variable**

| DMR | # CpGs | P-value | Max Effect | Overlapping Promoters |
| --- | --- | --- | --- | --- |
| chr12: 122019006-122019080 | 4 | 9.76E-03 | -1.78E-02 | KDM2B-005, KDM2B-001, KDM2B-006, KDM2B-201, KDM2B-004, KDM2B-007, KDM2B-008, RP13-941N14.1-001, KDM2B-002 |
| chr14: 55907122-55907501 | 8 | 5.62E-03 | 2.14E-02 | TBPL2-001 |
| chr15: 28147928-28148431 | 4 | 2.57E-03 | 2.12E-02 | NA |
| chr15: 98195808-98196247 | 4 | 1.37E-04 | -3.35E-02 | NA |
| chr16: 89686618-89687052 | 5 | 5.62E-03 | 1.97E-02 | DPEP1-002 |
| chr18: 67069959-67070461 | 6 | 6.10E-04 | -2.53E-02 | DOK6-001 |
| chr19: 18698825-18699631 | 9 | 1.37E-04 | -4.15E-02 | C19orf60-001, C19orf60-002, C19orf60-007, C19orf60-005, C19orf60-008, C19orf60-004, C19orf60-003, C19orf60-006 |
| chr2: 20211771-20211868 | 2 | 5.62E-03 | -1.32E-02 | MATN3-001, MATN3-201 |
| chr4: 298926-  299370 | 7 | 5.62E-03 | -1.46E-02 | ZNF732-001 |
| chr4: 74734714-74735092 | 8 | 5.62E-03 | -4.18E-03 | CXCL1-001, CXCL1-002 |
| chr6: 33282736-33283293 | 21 | 5.62E-03 | -1.60E-02 | TAPBP-008, TAPBP-001, TAPBP-003, TAPBP-209, TAPBP-010, TAPBP-007, TAPBP-004, TAPBP-002, TAPBP-006 |
| chr6: 90121670-90121836 | 2 | 5.62E-03 | -2.38E-03 | RRAGD-002, RRAGD-001, RRAGD-003 |
| chr6: 99395968-99396345 | 6 | 5.62E-03 | -7.98E-03 | FBXL4-201, FBXL4-001 |
| chr8: 142183507-142183677 | 3 | 6.29E-03 | 2.16E-02 | DENND3-019 |

**Table S4: DMR results for meta-analysis EPDS threshold-13 variable**

| DMRs | # CpGs | P-value | Max Effect | Overlapping Promoters |
| --- | --- | --- | --- | --- |
| chr1: 159046391-159047163 | 7 | 3.54E-03 | -3.46E-02 | AIM2-001 |
| chr1: 228646841-228647248 | 5 | 4.39E-03 | -4.81E-03 | HIST3H2A-001, HIST3H2BB-001 |
| chr1: 62660188-62660861 | 7 | 1.25E-06 | -2.93E-02 | L1TD1-001 |
| chr1: 99469323-99469698 | 4 | 3.54E-03 | 2.05E-02 | LPPR5-002, LPPR5-001, RP5-896L10.1-001, LPPR5-003 |
| chr10: 94820892-94821085 | 4 | 3.54E-03 | -1.38E-02 | CYP26C1-001, RP11-348J12.2-001 |
| chr11: 14993378-14994230 | 16 | 3.54E-03 | -1.22E-02 | CALCA-001, CALCA-201, CALCA-202, CALCA-003, CALCA-002, CALCA-004, CALCA-005 |
| chr12: 133022423-133022853 | 4 | 7.83E-03 | 4.77E-02 | NA |
| chr14: 24422956-24423618 | 5 | 3.54E-03 | -7.51E-03 | DHRS4-201, DHRS4-002, DHRS4-202, DHRS4-203, DHRS4-001, DHRS4-003, DHRS4-AS1-004, DHRS4-204, DHRS4-004, DHRS4-AS1-003, DHRS4-AS1-005, DHRS4-AS1-002, DHRS4-AS1-007, DHRS4-005, DHRS4-007, DHRS4-006 |
| chr16: 838502-838515 | 2 | 8.65E-03 | 1.53E-03 | RPUSD1-001, CHTF18-001, CHTF18-201, CHTF18-008, CHTF18-002, CHTF18-006, CHTF18-003, CHTF18-005, CHTF18-004, CHTF18-007, CHTF18-012, RPUSD1-004, RPUSD1-010, CHTF18-014, RPUSD1-009, RPUSD1-005, RPUSD1-006, RPUSD1-002, RPUSD1-003, RPUSD1-008, CHTF18-015, RPUSD1-007, CHTF18-013 |
| chr17: 46018875-46019184 | 6 | 4.28E-03 | 4.56E-03 | PNPO-001, AC003665.1-003, AC003665.1-002, PNPO-002, AC003665.1-001, PNPO-201, PNPO-202, PNPO-003, PNPO-004, PNPO-008, PNPO-005, AC003665.1-004, PNPO-007 |
| chr18: 67069959-67070461 | 6 | 3.62E-10 | -2.32E-02 | DOK6-001 |
| chr19: 51774377-51774666 | 5 | 4.05E-03 | 2.72E-02 | CTD-3187F8.11-003, CTD-3187F8.11-001, CTD-3187F8.2-001, CTD-3187F8.11-002 |
| chr19: 55598782-55599320 | 4 | 3.98E-03 | 3.93E-02 | EPS8L1-014, EPS8L1-019, EPS8L1-018 |
| chr19: 57306631-57307081 | 6 | 2.66E-03 | 1.86E-02 | NA |
| chr5: 1726145-1726243 | 3 | 4.05E-03 | 2.44E-02 | CTD-2587M23.1-001 |
| chr6: 151646312-151647133 | 10 | 7.79E-04 | 4.58E-02 | AKAP12-002 |
| chr6: 33282624-33283189 | 20 | 3.54E-03 | -1.46E-02 | TAPBP-008, TAPBP-001, TAPBP-003, TAPBP-209, TAPBP-010, TAPBP-007, TAPBP-004, TAPBP-002, TAPBP-006 |
| chr8: 22132563-22133356 | 11 | 3.54E-03 | 2.27E-02 | PIWIL2-001, PIWIL2-003, PIWIL2-002, CTD-2530N21.4-001 |
| chr8: 52321814-52322341 | 6 | 5.27E-03 | 3.13E-02 | PXDNL-004, PXDNL-003 |

**Table S5: DMR results for meta-analysis BDI-II threshold-14 variable**

| DMRs | # CpGs | P-value | Max Effect | Overlapping Promoters |
| --- | --- | --- | --- | --- |
| chr10: 2543763-2544120 | 2 | 2.63E-03 | 5.37E-02 | RP11-526P5.1-001, RP11-526P5.2-001, RP11-526P5.2-002 |
| chr10: 26942165-26942225 | 2 | 5.87E-03 | -7.70E-03 | NA |
| chr10: 70321874-70321889 | 2 | 7.42E-03 | 2.77E-02 | TET1-001 |
| chr11: 65190825-65191707 | 4 | 2.93E-06 | 6.40E-02 | NEAT1-002, NEAT1-001, NEAT1-202 |
| chr12: 104697193-104697983 | 12 | 1.00E-08 | 2.74E-02 | EID3-001 |
| chr13: 51417686-51418020 | 5 | 6.89E-03 | 1.93E-02 | DLEU7-002, DLEU7-001 |
| chr14: 105167300-105167457 | 2 | 6.22E-03 | 1.53E-02 | NA |
| chr17: 41738893-41739926 | 6 | 1.18E-04 | 7.00E-02 | MEOX1-001, MEOX1-201, MEOX1-003, MEOX1-002 |
| chr19: 49540073-49540241 | 3 | 9.74E-04 | 2.12E-02 | CGB1-001, CTB-60B18.6-001, CGB1-002, CTB-60B18.6-002 |
| chr20: 32856747-32857227 | 6 | 1.32E-06 | 2.74E-02 | NA |
| chr22: 24104820-24105692 | 6 | 1.50E-05 | -5.60E-02 | C22orf15-003, C22orf15-001, C22orf15-004, C22orf15-002, C22orf15-005 |
| chr3: 65342216-65342971 | 6 | 1.27E-04 | 2.65E-02 | NA |
| chr5: 145215546-145215784 | 3 | 9.74E-04 | 5.38E-02 | PRELID2-004, PRELID2-201, PRELID2-002, PRELID2-009, PRELID2-001, PRELID2-003, PRELID2-007 |
| chr5: 16508920-16509123 | 4 | 5.08E-03 | -2.11E-02 | FAM134B-003, FAM134B-006, FAM134B-004 |
| chr5: 191793-191806 | 2 | 7.64E-03 | 1.87E-02 | LRRC14B-001 |
| chr6: 32847530-32847845 | 13 | 4.05E-03 | 2.41E-02 | PPP1R2P1-002, PPP1R2P1-001 |
| chr6: 33047944-33048879 | 15 | 2.87E-07 | 4.45E-02 | HLA-DPB1-002, HLA-DPA1-004, HLA-DPA1-001, HLA-DPB1-008, RPL32P1-001, HLA-DPA1-002, HLA-DPB1-006, HLA-DPA1-005, HLA-DPB1-009, HLA-DPB1-005, HLA-DPB1-007 |
| chr7: 155174726-155175340 | 4 | 1.11E-04 | 3.26E-02 | AC008060.7-001 |
| chr7: 27183591-27185282 | 32 | 2.27E-04 | 2.51E-02 | HOXA5-001, HOXA-AS3-001, HOXA5-002, HOXA-AS3-005 |

**Table S6: DMR results for meta-analysis BDI-II threshold-20 variable**

| DMRs | # CpGs | P-value | Max Effect | Overlapping Promoters |
| --- | --- | --- | --- | --- |
| chr10: 70321668-70322442 | 5 | 4.18E-04 | 3.68E-02 | TET1-001 |
| chr11: 2020101-2020560 | 12 | 9.71E-04 | 1.43E-02 | H19-008, H19-004, H19-002, H19-001, H19-005, H19-009, H19-007, H19-003, H19-006 |
| chr13: 113622539-113622750 | 7 | 1.04E-03 | -1.69E-02 | MCF2L-202, MCF2L-002, MCF2L-AS1-001, MCF2L-005 |
| chr15: 70387217-70387268 | 2 | 4.25E-03 | 1.37E-02 | TLE3-010, TLE3-030, TLE3-022, TLE3-015, TLE3-026, TLE3-011, TLE3-008, TLE3-007, TLE3-013, TLE3-012, TLE3-020, TLE3-014 |
| chr17: 19290353-19290762 | 7 | 1.72E-03 | 6.60E-03 | MFAP4-001, MFAP4-002, MFAP4-003, MFAP4-004 |
| chr17: 37893764-37894636 | 10 | 2.76E-04 | 1.69E-02 | GRB7-001, GRB7-201, GRB7-009, GRB7-002, GRB7-006, GRB7-005, GRB7-011, GRB7-014, GRB7-012, GRB7-013, GRB7-008, GRB7-010 |
| chr18: 67069959-67070461 | 6 | 1.86E-07 | -2.41E-02 | DOK6-001 |
| chr2: 27530670-27531535 | 10 | 9.03E-07 | -2.31E-02 | UCN-001 |
| chr5: 1393934-1394633 | 7 | 4.05E-08 | 4.17E-02 | NA |
| chr6: 30458519-30458998 | 5 | 7.46E-04 | -1.44E-02 | HLA-E-001, HLA-E-002, HLA-E-003 |
| chr6: 33040535-33040610 | 2 | 6.84E-03 | 2.28E-02 | HLA-DPA1-205, HLA-DPA1-007, HLA-DPA1-003 |
| chr6: 33047944-33049360 | 16 | 4.05E-08 | 4.71E-02 | HLA-DPB1-002, HLA-DPA1-004, HLA-DPA1-001, HLA-DPB1-008, RPL32P1-001, HLA-DPA1-002, HLA-DPB1-006, HLA-DPA1-005, HLA-DPB1-009, HLA-DPB1-005, HLA-DPB1-007 |
| chr7: 155174726-155175340 | 4 | 1.98E-04 | 3.78E-02 | AC008060.7-001 |
| chr8: 1764878-1765820 | 12 | 1.29E-04 | -1.16E-02 | MIR596-201 |
| chr8: 70378380-70378994 | 7 | 1.19E-05 | 3.36E-02 | SULF1-201, SULF1-001, SULF1-008, SULF1-009, SULF1-010 |

**Table S7: Results for cg08667740 and cg22868225 for this study and the Viuff, A et al. study**

|  | Drakenstein Child Health Study^a^ | | Viuff, A. et al. ALSPAC mid-pregnancy depression^b^ | | Viuff, A. et al. Generation R Study^c^ | |
| --- | --- | --- | --- | --- | --- | --- |
| CpG sites | Effect | P-value^d^ | Effect | P-value | Effect | P-value |
| cg08667740 | -2.94E-04 | 0.755 | -0.025 | 3.90E-08 | 0.003 | 0.186 |
| cg22868225 | -3.28E-04 | 0.402 | -0.005 | 5.98E-08 | -0.001 | 0.672 |

a - in association with the EPDS 13 threshold depression variable

b - in association with the EPDS 12 threshold depression variable

c - in association with the Brief Symptom Inventory (BSI) 0.80 threshold variable

d - adjusted with Bacon and Cate

**Table S8: Results for cg06808585, cg05245515, and cg15264806 for this study and the Cardenas, A et al. study**

|  | Drakenstein Child Health Study^a^ | | Cardenas, A. et al. Discover cohort Project^a^ | | Cardenas, A. et al. Generation R Study^b^ | |
| --- | --- | --- | --- | --- | --- | --- |
| CpG sites | Effect | P-value^c^ | Effect | P-value^d^ | Effect | P-value |
| cg06808585 | -4.10E-03 | 0.359 | 3.10 | <0.05 | 0.04 | 0.96 |
| cg05245515 | 5.68E-03 | 0.177 | -1.59 | <0.05 | 0.28 | 0.29 |
| cg15264806 | -1.11E-04 | 0.609 | 0.14 | <0.05 | 0.05 | 0.63 |

a - in association with the EPDS 13 threshold depression variable

b - in association with the Brief Symptom Inventory (BSI) 0.80 threshold variable

c - adjusted with Bacon and Cate

d - FDR

**Table S9: Correlation between brain and blood DNAm for CpG sites and DMRs**

| DMR | CpG sites | DNAm Correlation Across Brain and Blood^a^ | P-value^a^ |
| --- | --- | --- | --- |
| N/A | cg04859497 | -0.184 | 0.422 |
| chr8:70378380-70378994 | cg15351186 | 0.194 | 0.399 |
|  | **cg07051728** | **0.499** | **0.023** |
|  | cg12181083 | 0.166 | 0.470 |
|  | cg04845579 | 0.216 | 0.346 |
|  | cg02283643 | 0.182 | 0.428 |
|  | cg07073960 | 0.201 | 0.380 |
|  | cg00613562 | -0.022 | 0.926 |
| chr11:65190825-65191707 | cg04145264 | 0.143 | 0.535 |
|  | cg09411730 | -0.418 | 0.060 |
|  | cg07985890 | -0.381 | 0.090 |
|  | cg18019132 | -0.168 | 0.466 |
| chr12:104697193-104697983 | cg01857475 | -0.131 | 0.570 |
|  | **cg09884423** | **0.543** | **0.012** |
|  | cg10572274 | -0.135 | 0.558 |
|  | cg18633684 | 0.226 | 0.323 |
|  | cg03817911 | 0.247 | 0.280 |
|  | cg20923245 | 0.243 | 0.287 |
|  | **cg27205904** | **0.677** | **0.001** |
|  | cg21234561 | 0.177 | 0.442 |
|  | cg26614816 | 0.370 | 0.099 |
|  | **cg09477407** | **0.552** | **0.011** |
|  | cg05057777 | -0.005 | 0.984 |
|  | cg01848457 | 0.345 | 0.125 |
| chr18:67069959-67070461 | **cg03790988** | **0.439** | **0.048** |
|  | cg20103519 | 0.160 | 0.487 |
|  | cg19513940 | 0.079 | 0.733 |
|  | cg00433861 | -0.177 | 0.442 |
|  | cg13890379 | -0.236 | 0.301 |
|  | cg20169576 | 0.079 | 0.733 |

^a^ - These values came from IMAGE-CpG

**Table S10: Sensitivity analysis results for the significant single CpG sites with and without HIV exposure as a covariate**

|  |  | Without HIV exposed^a^ | | With HIV exposed^a^ | |
| --- | --- | --- | --- | --- | --- |
| CpG sites | Depression Variable | Effect Δ beta^b^ | p-value | Effect Δ beta^b^ | p-value |
| cg22798925 | EPDS Continuous | 0.0011 | 1.06E-07 | 0.0011 | 7.27Ee-08 |
| cg04859497 | BDI-II Threshold 20 | -0.0642 | 8.09E-10 | -0.0645 | 6.89E-10 |
| cg27278221 | BDI-II Threshold 14 | -0.0195 | 5.40E-08 | -0.0194 | 6.16E-08 |

^a^ - All association models were adjusted for covariates: mother’s smoking status, average household income, child’s sex, preterm birth (<37 weeks), first three cell type PCs, and first five genotype PCs.

^b^ - Δ beta: This coefficient represents the mean difference of DNAm beta values between children of mothers who were screened positive for depression versus of those who were not. Negative coefficients refer to smaller mean DNAm beta values in children of mothers who were screened positive and positive coefficients refer to larger mean DNAm beta values in children of mothers who were screened positive for depression.
